# Conversational multi-turn interaction does not ensure triage-disposition alignment in ChatGPT Health in real and synthetic patient encounters

**DOI:** 10.64898/2026.07.21.26358588

**Authors:** Alvira Tyagi, Bennett Taylor, Pushkala Jayaraman, Mateen Jangda, Joy Jiang, Sanjana Kavula, Michael A. Gorin, Hannah Hugo, Nicholas Gavin, Robert Freeman, Alexander W. Charney, Ankit Sakhuja, Yuan Luo, Lisa Stump, Ashwin Ramaswamy, Girish N. Nadkarni, Bilal A. Naved

**Affiliations:** The Windreich Department of Artificial Intelligence and Human Health, Icahn School of Medicine at Mount Sinai, New York, NY, USA; Icahn School of Medicine at Mount Sinai, New York, NY, USA; The Milton and Carroll Petrie Department of Urology, Icahn School of Medicine at Mount Sinai and Mount Sinai Health System, New York, NY, USA; The Charles Bronfman Institute for Personalized Medicine, Icahn School of Medicine at Mount Sinai, New York, NY, USA; University of Miami Miller School of Medicine, Miami, FL, USA; Rice University, Houston, TX; Department of Medicine, NYC Health + Hospitals / Elmhurst, Icahn School of Medicine at Mount Sinai and Mount Sinai Health System, New York, NY, USA; Department of Emergency Medicine, Icahn School of Medicine at Mount Sinai, New York, NY, USA; Mount Sinai Health System, New York, NY, USA; The Hasso Plattner Institute for Digital Health at Mount Sinai, Icahn School of Medicine at Mount Sinai and Mount Sinai Health System, New York, NY, USA; Department of Preventive Medicine and Bioinformatics, Northwestern University Feinberg School of Medicine, Chicago, IL, USA

**Keywords:** artificial intelligence, patient safety

## Abstract

Individuals increasingly use conversational AI systems for symptom guidance. Whether multi-turn interactions improve clinical triage standard alignment remains uncertain. We conducted a retrospective, cross-sectional evaluation of 255 cases from three physician-reviewed sources: clinically authored vignettes (n=39), and real-world emergency department (N=76) and nurse line cases (n=140). CGPTH single-turn generated triage recommendations after only receiving an initial symptom description, reflecting potential typical use. Second, CGPTH multi-turn simulated nurse triage by asking subsequent questions before generating triage recommendations. Against nurse line standards, 52.9% single-turn and 55.7% multi-turn use prompting agreed exactly; clinician adjudicated disposition agreement was 54.1% and 48.2%. Discordant case recommendations represented lower acuity against nurse triage (natural: 70.8% under-triage, *P* < 0.0001; multi-turn: 69.0%, *P* < 0.0001). These findings suggest conversational interaction does not ensure safe triage-disposition alignment. Alongside aggregate agreement, clinical AI systems evaluations for symptom guidance should measure ordinal distance from standards, error direction, and additional dialogue conditions that affect recommended care standards.

## INTRODUCTION

Patients increasingly use conversational artificial intelligence (AI) systems for guidance about new or concerning symptoms, including questions about whether and how urgently to seek care.^1–3^ Although consumer systems such as ChatGPT Health are positioned to support medical care, their responses influence whether a person stays home, contacts a clinician, seeks urgent care or presents to an emergency department. When symptom guidance includes advice about next steps, it becomes a triage-relevant disposition problem: how urgently should this person seek care? Clinical triage maps incomplete patient-reported information to an urgency level under uncertainty, balancing the risk of missed illness against the burden of unnecessary utilization. The direction of error matters: over-triage increases avoidable healthcare use, whereas under-triage delays needed evaluation. The central question is whether conversational interaction helps patient-facing large language models (LLMs) align with established clinical triage standards.

Prior studies of digital symptom checkers and LLMs have found widely variable triage performance, with estimates depending on the platform, clinical presentation, and testing format.^1,4–7^ Evaluations have used simulated vignettes, single-turn inputs, or structured prompts, leaving open the possibility that static testing artificially underestimates systems designed for interactive dialogue.^2,5^

However, more conversation does not presuppose better clinical triage. While structured nurse-triage protocols utilize algorithmic, sequential branching logic to systematically converge on a definitive disposition, conversational AI typically generates flexible, free-text guidance that may not reliably map to clinician- or protocol-derived urgency levels. Taken together, these limitations leave three central uncertainties regarding patient-facing use: whether multi-turn interaction improves disposition-level alignment, whether model performance generalizes from synthetic vignettes to real-world clinical encounters, and whether discordant AI recommendations are directionally balanced versus systematically biased toward dangerous under-triage or unnecessary over-triage.

We therefore evaluated ChatGPT Health in 255 clinical triage cases spanning standardized patient-language vignettes, real-world emergency department (ED) encounters adjudicated by attending emergency physicians, and real-world nurse-line encounters adjudicated by a health-system medical director with contemporaneous Schmitt-Thompson protocol recommendations.^8^ This design allowed us to test whether conversational interaction improves triage-disposition alignment across controlled vignettes and real-world encounter-derived cases, while rigorously quantifying the magnitude and direction of disagreement when AI guidance diverged from clinical reference standards.

## RESULTS

### Agreement with reference standards

**Fig 1**. Across 255 adjudicated cases, CGPTH demonstrated similar performance under the natural and multi-turn interaction paradigms (Fig. 1).

**FIGURE 1.**
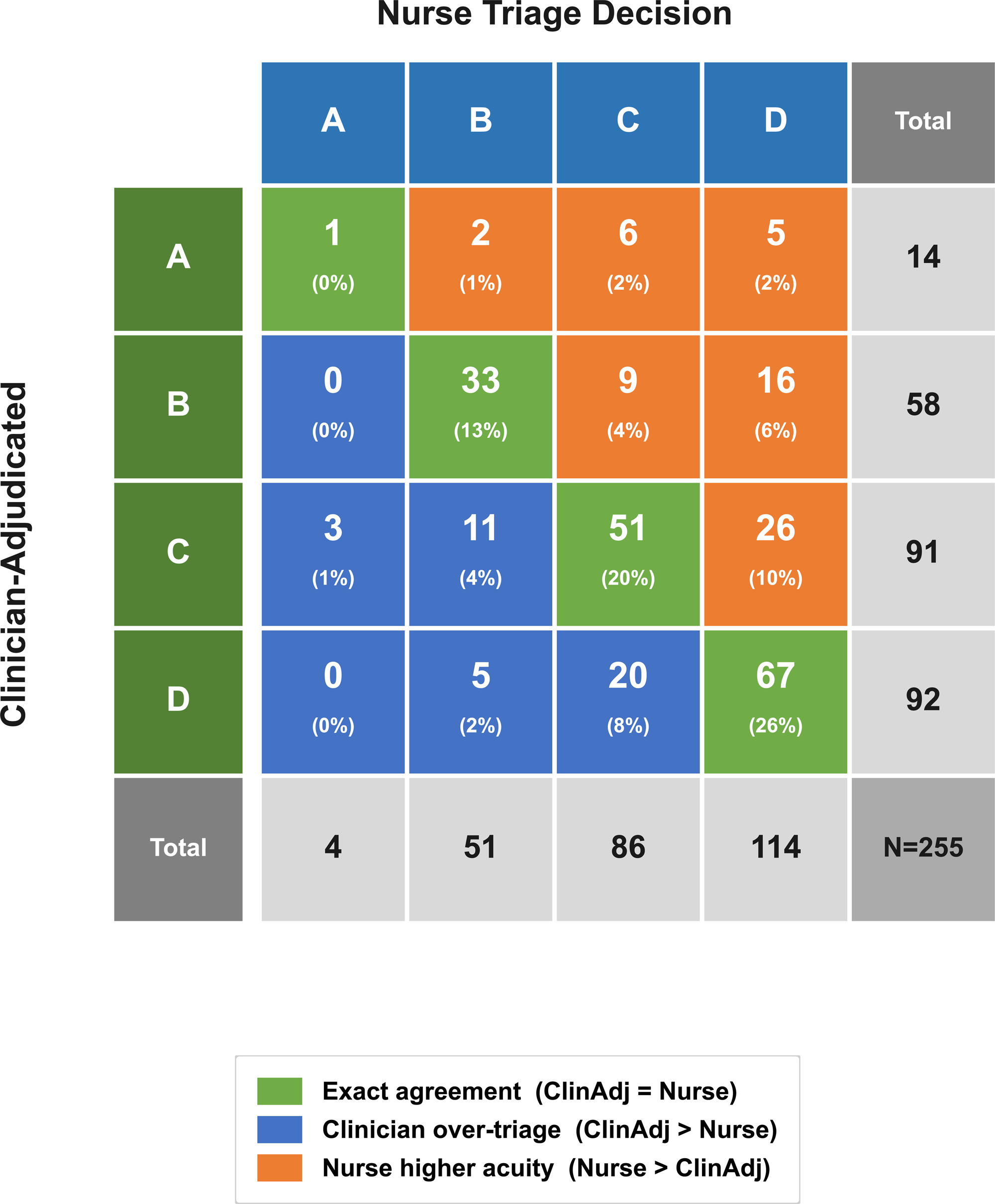
ChatGPT Health triage errors by use condition and reference standard. From left to right, Clinician-Authored Vignettes, Emergency Department and Nurse Triage are depicted for the natural condition (bottom) and multi-turn condition (top), with segmented bar graphs for under-triage (orange), concordance (gray), and over-triage (blue). Counts are listed beneath each percentage in parentheses. Against nurse triage, disagreement skewed disproportionally towards under triage, with 70.8% in the natural condition (95% CI 61.8–78.8, P < 0.0001) and 69.0% in the multi-turn condition (59.6–77.4, P < 0.0001). Safety critical (≥ 2 levels below reference) under-triage rates increased from the synthetic vignettes dataset (5.1%) to the human rater datasets (10.5% and 17.9% for ED and nurse triage respectively), demonstrating a tendency to under-triage more often in real-world presentations.

Compared with the clinician-adjudication reference standard, ChatGPT Health achieved exact agreement in 54.1% of single turn cases (95% CI 47.8–60.4%; weighted Cohen’s κ = 0.44, 95% CI 0.35–0.51) and 48.2% of multi-turn cases (95% CI 42.0–54.6%; κ = 0.36, 95% CI 0.28–0.44) (Fig. 1). Mean ordinal distance from the clinician standard was 0.59 (SD 0.73) for natural use and 0.69 (SD 0.78) for multi-turn prompting. Where ChatGPT Health and clinician adjudicated standards disagreed, 60.7% of natural-use disagreements represented under-triage (95% CI 51.2–69.6%) and 39.3% represented over-triage (95% CI 30.4–48.8%; two-sided binomial *P* = 0.026).

In multi-turn prompting, 54.5% of discordant cases represented under-triage (95% CI 45.7–63.2%) and 45.5% represented over-triage (95% CI 36.8–54.3%; *P* = 0.34). Among errors, 73.5% (n = 86 of 117, 95% CI 64.5–81.2%) of natural-use cases and 72.7% (n = 96 of 132, 95% CI 64.3–80.1%) of multi-turn cases differed by a single adjacent triage category; however, 26.5% (n = 31 of 117, 95% CI 18.8–35.5%) and 27.3% (n = 36 of 132, 95% CI 19.9–35.7%) of disagreements, respectively, deviated by two or more triage categories (for example, recommending care within 2 days versus immediate ED evaluation). Safety-critical under-triage occurred in 18 of 255 natural-use cases (7.1%, 95% CI 4.2–10.9%) and 17 of 255 multi-turn cases (6.7%, 95% CI 3.9–10.5%). Over-triage was most prevalent for multi-turn use compared to the clinician-adjudicated reference standard (60/255, 23.5%; 95% CI, 18.5–29.2%). Utilization-critical over-triage was also greatest for multi-turn versus clinician adjudication (19/255, 7.5%; 95% CI, 4.5–11.4%).

**Fig 2**. To determine the severity of discordant responses, we examined the percentage of all 255 cases at each ordinal distance (0 = exact agreement, 1-3 disagreement levels) for the natural condition (blue) and multi-turn (orange) across each reference condition (Fig. 2).

**FIGURE 2.**
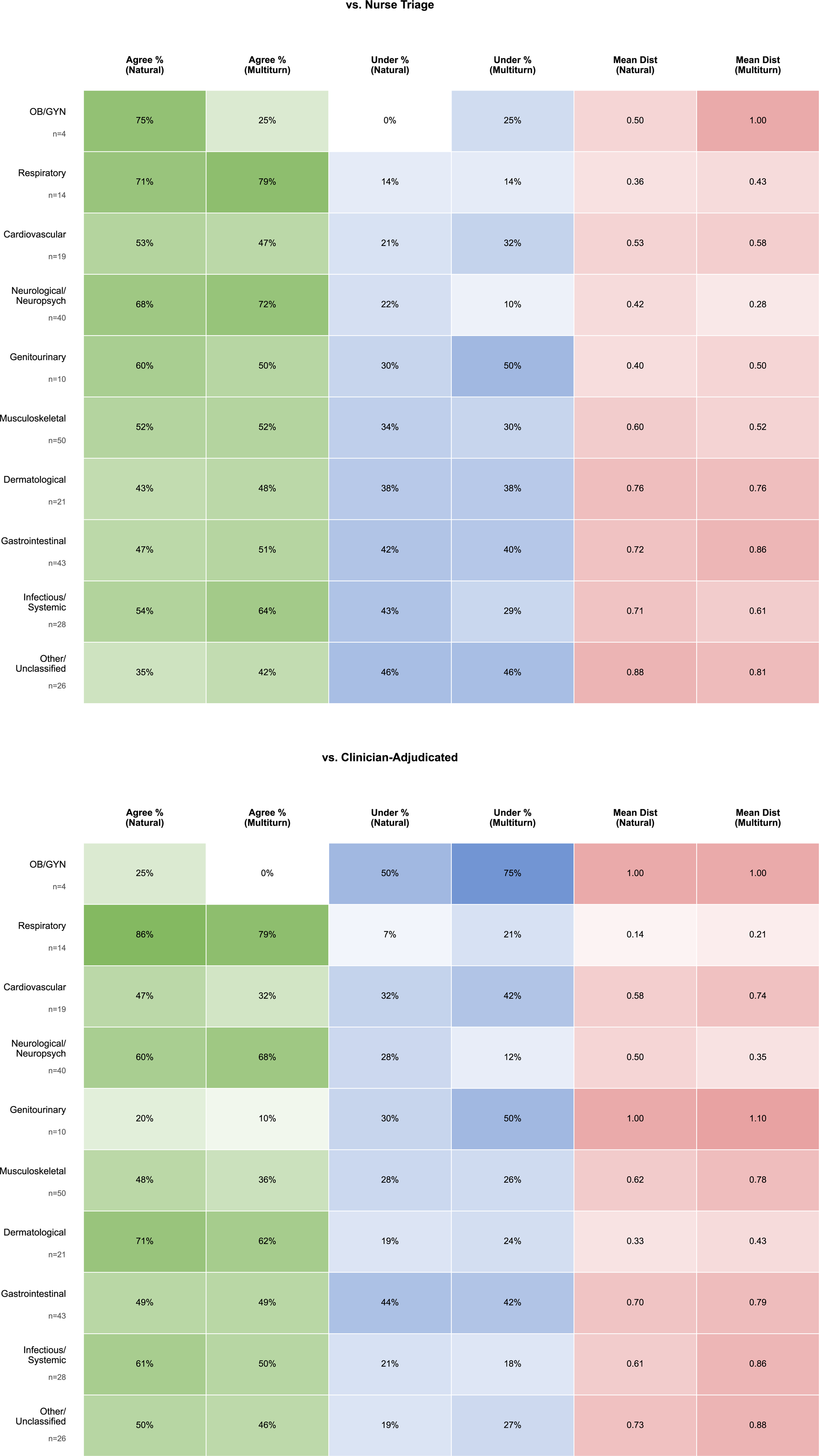
Clinical Distance distribution between ChatGPT Health and reference standards. Clinical distance is the absolute ordinal distance (0-3 levels, A-D scale) between per-case recommendation and reference. Each bar depicts total case (*n* =255) percentage at each difference for natural (blue) and multi-turn (orange) use conditions against clinician adjudicated (A) and nurse triage (B) reference standards. Most cases grouped at 0-1 levels, with only moderate ordinal agreement (weighted κ: natural 0.437 [95% CI 0.351–0.514] and multi-turn 0.364 [0.281–0.441] vs clinician-adjudicated; natural 0.395 [0.316–0.474] and multi-turn 0.418 [0.338–0.499] vs nurse triage). ≥ 2 levels of disagreement were seen in 11.7-14.1% of cases, with mean distance distribution not differing significantly against clinician adjudication after Bonferroni correction (multi-turn mean distance 0.686 vs natural 0.592; Wilcoxon *P =* 0.029). This shows that additional conversation did not improve agreement with the clinician standard.

Compared with nurse triage recommendations, ChatGPT Health achieved exact agreement in 52.9% of natural-use cases (95% CI 46.6–59.2%; κ = 0.40, 95% CI 0.32–0.47) and 55.7% of multi-turn cases (95% CI 49.4–61.9%; κ = 0.42, 95% CI 0.34–0.50) (Fig. 1). Mean ordinal distance from nurse triage was 0.62 (s.d. 0.77) for natural use and 0.60 (s.d. 0.80) for multi-turn prompting. Where ChatGPT Health and nursing standards disagreed, 70.8% of natural-use disagreements represented under-triage (95% CI 61.8–78.8%) and 29.2% represented over-triage (95% CI 21.2–38.2%; two-sided binomial *P* < 0.0001; full classification for both standards in Supplemental Fig. 1).

This pattern persisted with multi-turn prompting, in which 69.0% of discordant cases represented under-triage (95% CI 59.6–77.4%) and 31.0% represented over-triage (95% CI 22.6–40.4%; P < 0.0001). Among errors, 75.0% of natural-use cases (n = 90 of 120, 95% CI 66.3–82.5%) and 71.7% of multi-turn cases (n = 81 of 113, 95% CI 62.4–79.8%) differed by a single adjacent triage category; however, 25.0% (n = 30 of 120, 95% CI 17.5–33.7%) and 28.3% (n = 32 of 113, 95% CI 20.2–37.6%) of disagreements, respectively, deviated by two or more triage categories. Safety-critical under-triage (two or more categories below nurse triage) occurred in 26 of 255 natural-use cases (10.2%, 95% CI 6.8–14.6%) and 22 of 255 multi-turn cases (8.6%, 95% CI 5.5–12.8%). Over-triage rates against the nurse-triage reference standard were equivalent between conditions overall (35/255, 13.7%; 95% CI, 9.8– 18.6% for both natural and multi-turn use). However, utilization-critical over-triage was more than twice as frequent for multi-turn compared to natural use (10/255, 3.9%; 95% CI, 1.9–7.1% vs. 4/255, 1.6%; 95% CI, 0.4–4.0%). Errors were acuity dependent, with under-triage concentrated at high acuity reference standards (C-D) and over-triage apparent in low acuity levels (A-B) (Supplemental Fig. 3).

**Fig 3**. Given the use of both nurse triage and clinician adjudication as reference standards, we next evaluated agreement between these benchmarks directly (Fig. 3).

**FIGURE 3.**
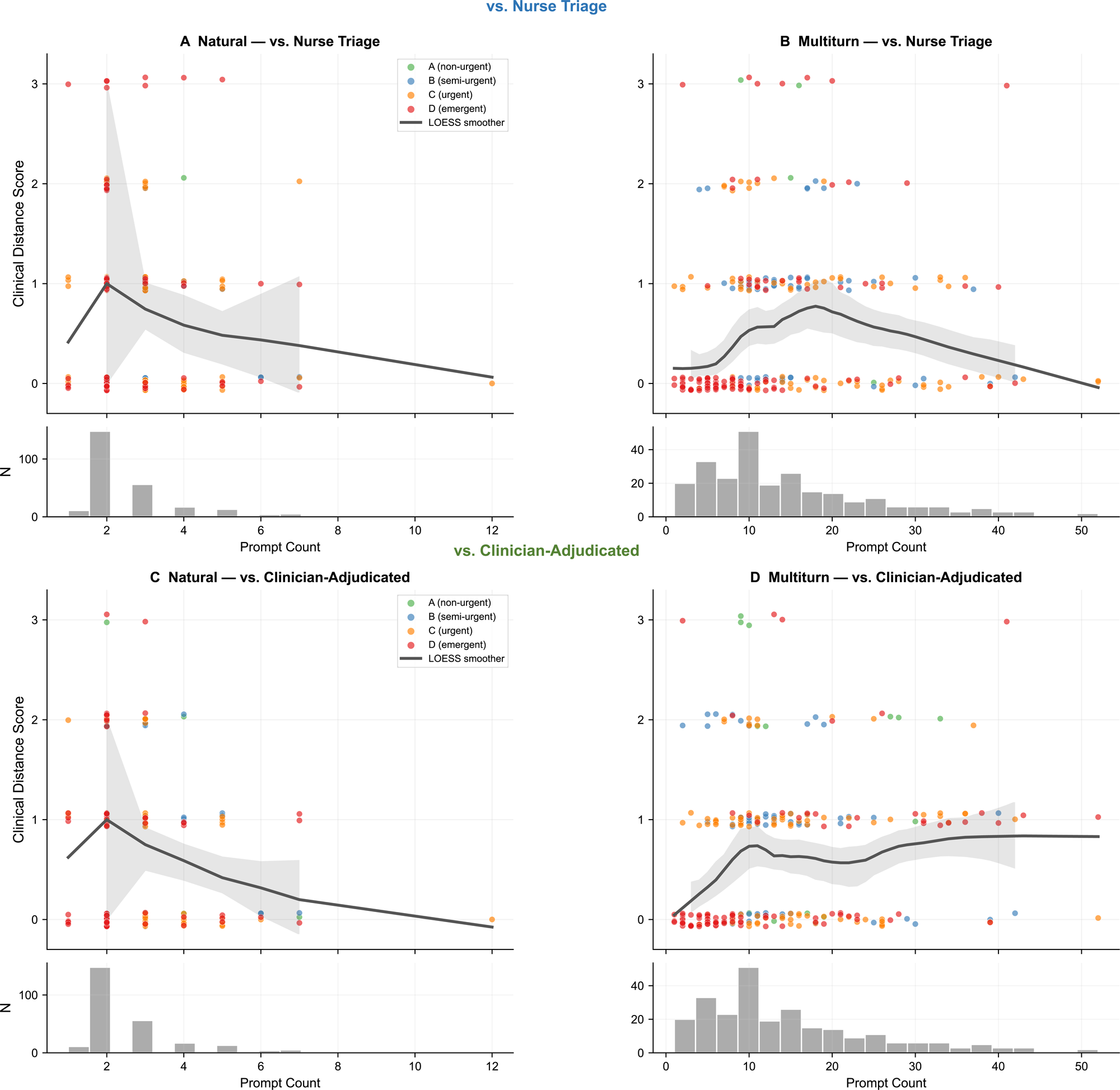
Concordance of clinician adjudicated and nurse triage reference standards. Bar graph shows percentage of total cases (*n* =255) by levels of concordance between the two reference standards. The standards agreed in 59.6% of cases (152/255; 95% CI 53.3–65.7), differed in one level in 26.7%, two levels in 11.8%, and the maximum three levels in only 5 of 255 cases (2.0%). Among 103 discordant cases, nurse triage held higher-acuity in 64 (62.1%) compared to clinician-adjudication 39 (binomial *P =* 0.018).

The clinician- and nurse-triage reference standards agreed in 152 of 255 cases (59.6%, 95% CI 53.3–65.7%), with 5 of 255 cases (2.0%, 95% CI 0.6–4.5%) differing by three triage categories (Fig. 3). Concordant recommendations occurred most frequently in higher-acuity categories (118 of 152 concordant pairs, 77.6%, 95% CI 70.2–84.0%). When the standards disagreed, nurse triage recommendations were more often higher acuity than clinician-triage recommendations (64 of 103, 62.1%, 95% CI 52.0–71.5%; two-sided binomial *P* = 0.018; full classification in Supplemental Fig. 6), a pattern that persisted among urgent and emergent presentations (62 of 101, 61.4%, 95% CI 51.2–70.9*%; P* = 0.028).

### Effect of Multi-Turn Interaction

Across the two interaction conditions, exact agreement with nurse triage did not differ significantly (McNemar’s χ² = 0.63*, P* = 0.43; Wilcoxon signed-rank test for ordinal distance, *r =* 0.02, *P* = 0.74). Against the clinician standard, multi-turn prompting showed a non-significant trend toward greater ordinal distance from the clinician standard (Wilcoxon *r* = 0.14, raw *P* = 0.029, Bonferroni-adjusted *P* = 0.059), with a non-significant trend toward lower exact agreement (McNemar’s χ² = 3.32, *P* = 0.068).

### Generalization across Data Sources

Performance generalized unevenly across data sources. Against nurse triage, exact agreement was statistically indistinguishable among the three datasets (natural, 59.0% / 60.5% / 47.1%; multi-turn, 53.8% / 57.9% / 55.0% for vignettes / ED / nurse-line; χ² *P* ≥ 0.12), and between synthetic vignettes and pooled real-world encounters (*P* ≥ 0.52). Against the clinician adjudicated standard, however, agreement was substantially lower for synthetic vignettes than for real-world encounters (natural, 30.8% vs 58.3%, χ² *P* = 0.0027, Mann–Whitney *P* = 0.0008; multi-turn, 30.8% vs 51.4%, *P* = 0.028), with a significant difference across all three datasets (natural, *P* = 0.0064). Thus, alignment with nurse-triage dispositions was stable from synthetic to real-world cases while physician-adjudicated dispositions did not fully generalize.

### Sensitivity and Reliability

Sensitivity analyses using conservative (favoring higher-acuity dispositions) and aggressive (favoring lower-acuity dispositions) adjudication approaches yielded stable exact agreement rates across all conditions (natural use, 49.8–55.7%; multi-turn, 55.7–58.4% across primary, conservative and aggressive resolution, Supplementary Fig. 2). Per-grader weighted κ was fair to moderate (0.13–0.51 vs nurse triage; 0.00–0.63 vs clinician adjudicated); one grader (Grader 3) fell >1 s.d. below the group mean against nurse triage; excluding that grader maintained primary directional findings (under-triage 65.3%/64.8%, both *P* < 0.01; Supplementary Table 1). Grader 5 was flagged against clinician adjudicated but only completed 14 synthetic vignette cases.

### Performance by Complaint and Conversation Length

Triage alignment varied by presenting complaint category (Supplementary Fig. 5). Respiratory (75.0% exact agreement) and neurological or neuropsychiatric (70.0%) complaints demonstrated the highest alignment with clinical standards, whereas unclassified (38.5%), dermatologic (45.2%) and gastrointestinal (48.8%) complaints exhibited lower exact agreement and greater mean ordinal clinical distance (0.85, 0.76 and 0.79 steps, respectively). Mean ordinal distance is defined as the number of triage steps between two ratings on the A-D scale; levels A and B are one step apart, whereas A and D are three steps apart. In the multi-turn condition, more turns were significantly associated with greater clinical distance from the reference standards (Spearman ρ = 0.162, n = 255, 95% CI 0.040–0.279, *P* = 0.010) (**Fig. 4**, **Supplementary Figure 4**) significant only in the nurse-line dataset under multi-turn, ρ = 0.27, *P* = 0.001). This association was not observed under the natural condition (ρ = 0.048, n = 255, 95% CI −0.08 to 0.17, *P* = 0.44). Thus, longer conversations did not correspond to reduced separation between ChatGPT Health recommendations and established reference standards.

**FIGURE 4.**
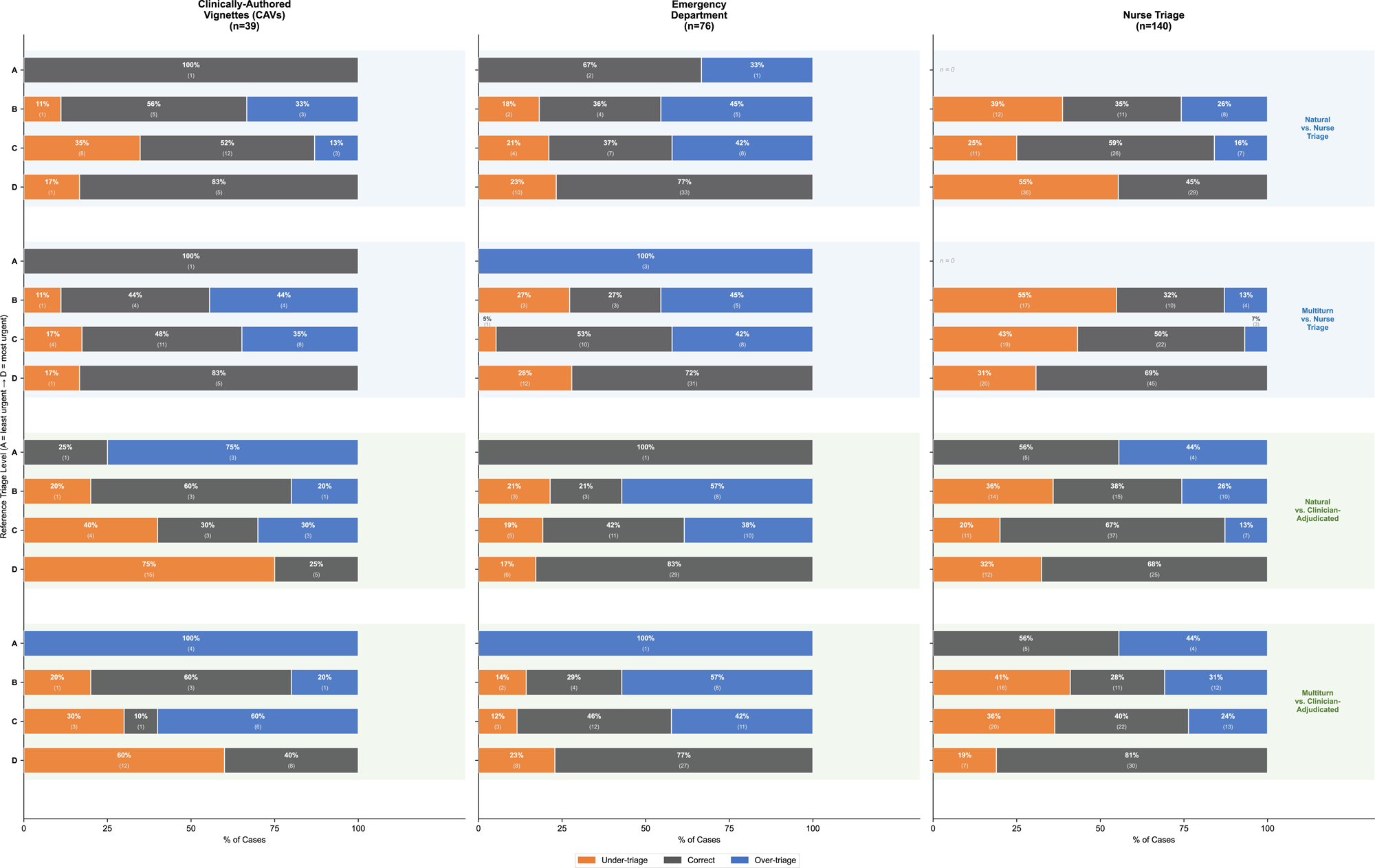
Multi-turn condition association of conversation length and clinical distance against nurse triage reference standard. Each point on the graph represents one case, plotted on the x-axis at total prompts before triage recommendation, and on the y-axis at clinical distance from the nurse triage reference standard with a LOESS smoother and 95% band. More prompts were significantly associated with greater distance (Spearman ρ = 0.162, n = 255, 95% CI 0.040–0.279, *P* = 0.010). Longer conversations therefore did not reduce separation from the reference disposition.

## DISCUSSION

In this evaluation of 255 clinical triage encounters spanning standardized vignettes, real-world emergency department encounters, and real-world nurse-line encounters, ChatGPT Health showed only moderate triage-disposition alignment with clinician adjudicated and nurse-triage reference standards. Exact agreement occurred in approximately half of cases under both natural-use and protocol-guided multi-turn conditions. Requiring sequential interaction did not consistently improve alignment: performance increased relative to nurse triage recommendations but decreased relative to clinician adjudicated dispositions (neither significant). Longer conversations did not correspond to less discordance from reference standards. The most safety-relevant finding was the direction of disagreement. When ChatGPT Health diverged from nurse triage recommendations, approximately 70% of discordant recommendations were lower acuity than the reference standard. These findings indicate that conversational interaction alone does not ensure triage-disposition alignment and that aggregate agreement measures can obscure clinically important directional error.

These results directly inform ongoing debates regarding how LLMs should be tested and deployed in healthcare.^9–11^ Prior evaluations of digital symptom checkers have frequently relied on static, single-turn prompts, prompting criticism that such testing artificially underestimates systems designed for interactive dialogue.^4^ However, our evaluation demonstrated that enabling multi-turn questioning did not reliably rescue triage performance; in fact, longer conversations were significantly associated with greater clinical distance from the correct disposition. This research challenges the prevailing assumption that conversational interactivity inherently improves clinical reasoning, demonstrating instead that open-ended generative dialogue cannot seamlessly replicate the sequential, algorithmic branching logic utilized by structured clinical triage protocols.

Furthermore, these findings highlight the critical importance of evaluating the directionality of algorithmic errors rather than relying solely on aggregate accuracy metrics. Clinical triage is fundamentally an exercise in decision-making under uncertainty, designed to balance the operational burden of unnecessary utilization against the direct risk of patient harm. While previous literature has documented the variable diagnostic accuracy of AI symptom checkers, less attention has been paid to the asymmetric clinical risks of their failures.^9–11^ Even accounting for baseline disagreement between human reference standards (which achieved exact agreement in 59.6% of cases) we observed a persistent under-triage bias relative to nurse triage. This study extends prior work by shifting the evaluative focus from absolute categorical accuracy to directional clinical safety, revealing that patient-facing generative AI systems exhibit a systematic propensity to provide false reassurance in acute medical scenarios.

Several limitations should be considered when interpreting these results, spanning chance, bias, and confounding. First, regarding chance, our exploratory subgroup analyses by chief complaint (e.g., obstetric and gynecologic presentations) contained small sample sizes, which limits statistical power and the ability to draw definitive conclusions about domain-specific performance. However, our primary aggregate endpoints were robustly powered, yielding highly significant findings for directional bias across the entire cohort. Second, regarding measurement bias, this study relied on standardized vignettes and retrospective encounter notes rather than live, prospective interactions. While real-world users might interact with the system erratically, utilizing adjudicated texts ensured a strict, reproducible gold standard, thereby isolating the model’s native triage reasoning from user-input variability. Finally, regarding confounding, the observed association between higher prompt counts and triage inaccuracy is likely confounded by underlying case complexity; inherently difficult clinical cases naturally elicit longer conversations and are simultaneously more prone to triage error. While this limits causal claims about conversational length, it confirms that the AI system fails to safely navigate the precise, complex scenarios where patients most require reliable guidance.

The systemic under-triage observed in this study carries urgent and specific implications across the healthcare ecosystem. For patients, the tendency of conversational AI to underestimate clinical severity means these platforms pose a severe risk of delayed care, necessitating the implementation of explicit, unavoidable user-interface warnings regarding acute symptoms. For providers, clinicians must routinely incorporate questions about prior AI consultation into their history-taking, as patients arriving with AI-generated false reassurance may require active de-anchoring from incorrect medical advice. For payers and health systems seeking to optimize healthcare utilization, replacing protocol-driven digital front doors with pure generative AI risks shifting the burden from manageable over-triage to costly, delayed acute care; payers must therefore integrate deterministic, algorithmic safety logic alongside conversational interfaces. Finally, for policymakers, the reality that consumer AI functions as a de facto triage tool necessitates regulatory action.

Agencies overseeing healthcare technologies should evaluate patient-facing triage language models under Software as a Medical Device framework, establishing and enforcing strict premarket safety thresholds against systemic under-triage before permitting widespread public deployment.

## METHODS

### Study Design

We conducted a retrospective, cross-sectional evaluation of ChatGPT Health (CGPTH), a conversational large language model (LLM)-based clinical assistant, across two experimental conditions: a natural conversational condition and a structured multi-turn conversational (termed “multiturn”) prompt condition. Both conditions were evaluated against nurse triage and clinician reference standards. This study did not require IRB review, as it involved exclusively de-identified, retrospectively derived clinical data and standardized clinical vignettes, and did not constitute human subjects research. The real-world emergency department and nurse line datasets are reported in accordance with the Strengthening the Reporting of Observational Studies in Epidemiology (STROBE) guidelines for observational studies (Supplementary Table 2).

### Data Source and Case Selection

The evaluation dataset comprised three separate datasets for a total of 255 cases (Table 1, Extended Data Fig. 1; full prompt listing in Extended Data Table 1). The first consisted of 39 standardized clinical vignettes from Ramaswamy et al. 2026, spanning 21 clinical domains.^4^ The full vignette set was used without further selection. Each vignette consisted of a single-sentence subjective complaint corresponding to a discrete clinical scenario with intentionally limited contextual information at the outset, reflecting how patients in each vignette may search their symptom. ^3^ The second was 76 real-world encounters from a community Emergency Department, where clinician decisions were determined by individual EM clinicians rating triage. Encounters were included if clinical documentation was complete, and a clinician triage disposition was available. All 76 encounters provided for this dataset met the criteria and were used in full. The third dataset comprised 140 encounter notes from a nurse telephone triage service, each independently reviewed by a medical director, who assigned a triage recommendation. Of 141 available cases, one case was a repeat and excluded from analysis. Each case consisted of a chief complaint and associated clinical context to generate a triage recommendation.

**TABLE 1.**
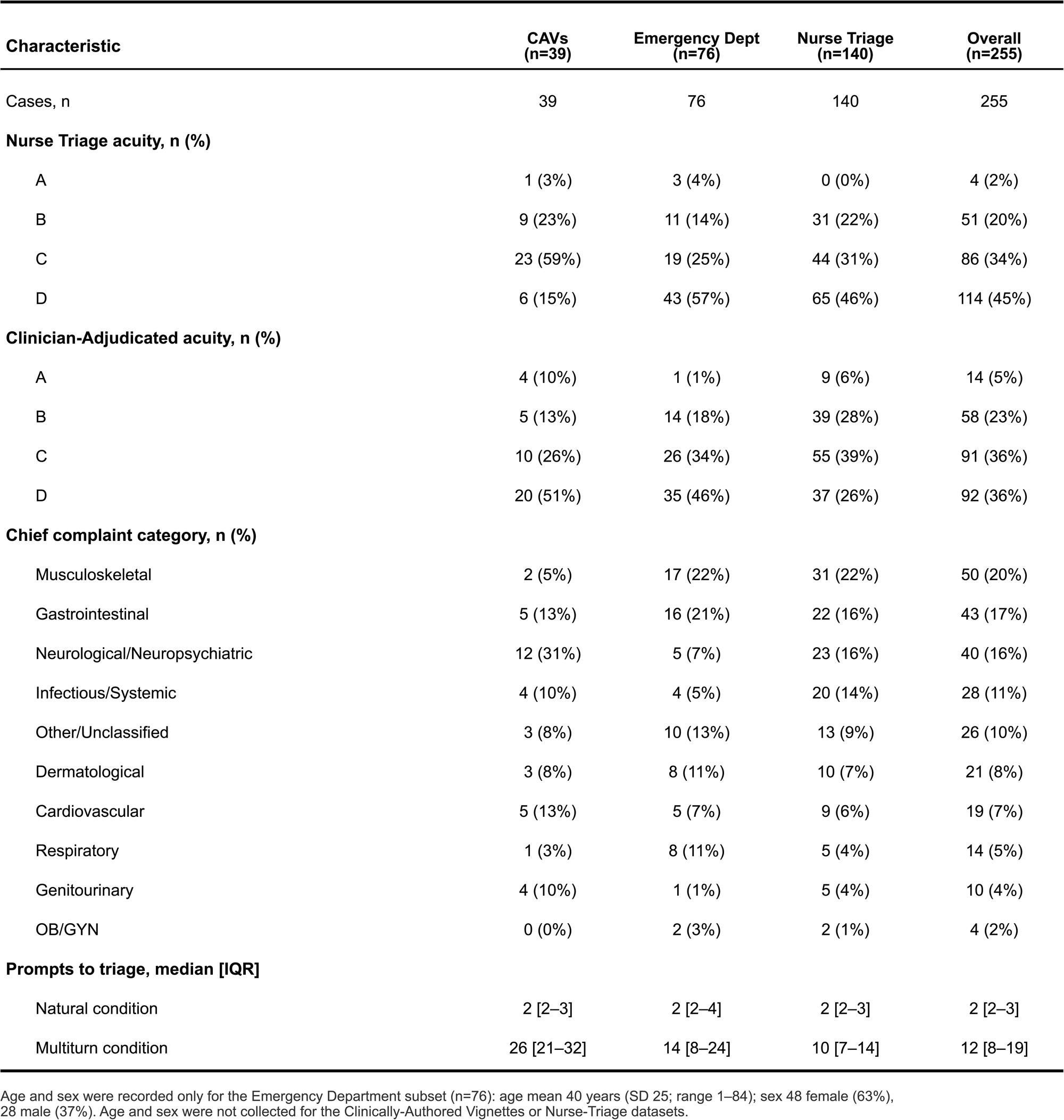
Case Characteristics by dataset. Summary of the 255 triage cases across all three datasets (Clinically-Authored Vignettes, *n* =39; Emergency Department, *n =*76; Nurse Triage, *n*= 140). Reference-standard triage distribution (A-D), chief complaint categories and prompt counts are given *n* (%) with each dataset. Age and sex were only included for Emergency Department dataset, noted in the footnote.

Nurse-triage reference dispositions were derived for all three datasets by graders applying Schmitt-Thompson protocols, a widely used standard for telephone triage; for the nurse-line dataset, these additionally reflected the encounters’ contemporaneous protocol-based dispositions.^1,3^ Encounter dates for emergency department and nurse line datasets were not kept for de-identification and not reported, while all ChatGPT Health evaluations occurred between March 5-May 21, 2026 (see LLM Conditions). No a priori power calculation was performed. Sample size was determined by available cases across three data sources.

### Triage Scale

Grading of triage corresponded to an A-D scale, where A represented home care, B to seek care with a provider over 24 hours, C to seek care within 24 hours, and D to go to the Emergency Room. Levels were treated as ordinal, with linear (equal-interval) spacing applied between adjacent levels in weighted analyses. Level D represents the highest acuity, and under-triage was designated the safety-critical error direction.

### LLM Conditions

CGPTH was evaluated as a representative conversational clinical AI system. The system is explicitly positioned as supplementary to, rather than a replacement for, professional medical care. In the natural condition, each case was presented by a single sentence with the chief complaint, with conversation continuing until the first explicit triage recommendation was given. In the multiturn condition, the same cases were presented, but CGPTH was instructed to ask one question at a time to arrive at a triage decision. When CGPTH requested additional information in either condition, graders answered one question at a time using details available in the case (the vignette text in Dataset 1, or the documented clinical details in Datasets 2 and 3). Information not contained in the case was answered as “I don’t know,” except where a question had an unambiguous answer given the case. When pain severity was not documented, it was recorded as moderate (4–6 of 10). All evaluations were conducted between March 5–May 21, 2026. Graders accessed the system through identical interface conditions using the standard consumer-facing application without custom system prompts or configuration modifications. Graders cleared system history to avoid prior case exposure.

### Grading

The two reference standards were clinician adjudication and nurse triage protocol determinations. Physician adjudication was recorded for each case, reflecting the decisions of either joint consensus of New York City physicians (A.R., H.H. and M.A.G., Dataset 1), an individual clinician (of 3 Emergency Medicine physicians at a community hospital in Maryland – N.M., S.A., or M.M., Dataset 2) or a system medical director of a large (>40) hospital system in the American South (E.E., Dataset 3). Nurse triage level decisions were determined for Dataset 1 and 2 by the graders, while already determined for Dataset 3. Physician adjudication composition varied across datasets, but all applied the same four level triage scale and definitions (see Triage Scale). Cases with range-valued triage decisions were treated as consensus across the range.

Four graders independently reviewed CGPTH outputs and assigned triage levels. Each grader assessed non-overlapping sets of prompts within each dataset, precluding direct pairwise inter-grader comparison among primary graders. Graders were blinded to reference-standard disposition at the time of assigning CGPTH triage levels. A fifth grader (Grader 5) served primarily as an independent reviewer, blinded to primary grader’s assignments during initial evaluation. Grader 5 was subsequently unblinded for reliability analyses. Grader 5 also completed 14 primary assessments in dataset 1, with this set flagged and reported separately in reliability analyses. Graders 2 (n = 62), 3 (n = 43) and 5 (n = 14) were medical students, 4 (n = 58) was an undergraduate, and 1 (n = 78) a postdoctoral fellow.

### Outcome Measures

The primary outcomes examined performance of both CGPTH conditions versus nurse-triage and clinician adjudicated reference standards, including exact agreement rate, Cohen’s weighted kappa, under and over triage rates with direction-of-error asymmetry, mean clinical distance score, safety-critical under-triage (ordinal distance ≥2 below reference standard) and utilization-critical over-triage (ordinal distance ≥2 from reference standard). Secondary outcomes analyzed sensitivity results for discordant triage decisions between the primary grader and reviewer in both a conservative (higher level selected if disagreement) resolution and aggressive (lower level selected) resolution condition. Other secondary outcomes included subgroup performance by dataset, chief complaint, and association of prompt count and accuracy.

### Scoring and Outcomes

A clinical distance score was defined as the ordinal difference between the assigned triage level and the reference standard (1 = minor, 2 = moderate, 3 = severe). For the 17 range-valued clinician adjudicated dispositions, all values within the range were considered as the reference. Under-triage was the pre-specified safety-critical error direction, and under-triage by ≥2 ordinal levels above reference standard was designated safety-critical. When the primary grader and independent reviewer assigned different triage levels, a sensitivity analysis applied three resolution strategies: primary, conservative (higher-acuity level selected) and aggressive (lower-acuity level selected); primary analyses used the primary strategy.

### Statistics

Analyses were performed in Python 3 (NumPy, SciPy and statsmodels; figures in Matplotlib; analysis assisted by Claude Code, Anthropic). All tests were two-sided with α = 0.05, except the two primary LLM-versus-reference-standard comparisons (nurse triage and clinician adjudicated, for which a Bonferroni-corrected α = 0.025 was applied). Ninety-five percent confidence intervals (CIs) are reported for all primary estimates.

Exact agreement was defined as cases with a clinical distance of 0. Exact agreement and all other proportions (direction-of-error, single-step and ≥2-step disagreement, and safety-critical rates) are reported with exact (Clopper–Pearson) binomial 95% CIs. Ordinal agreement was summarized with Cohen’s weighted κ (linear weights), with 95% CIs from 1,000 bootstrap resamples; mean clinical distance is reported with its standard deviation.

Agreement between clinician adjudicated and nurse triage reference standards was summarized by exact concordance with 95% CI, and directional disagreements were tested with a two-sided exact binomial test.

LLM performance was summarized for each interaction condition by the exact agreement rate (exact 95% CI), Cohen’s weighted κ, mean ordinal distance, and the proportion of under- and over-triage. The pre-specified primary safety analysis used a two-sided exact binomial test of the null hypothesis that under- and over-triage were equally likely among discordant cases. The proportion of errors classified as under-triage is reported with its exact 95% CI.

The natural and multi-turn conditions were compared on the same cases using McNemar’s test for exact agreement and the Wilcoxon signed-rank test for ordinal distance, evaluated separately against each reference standard and Bonferroni-corrected across the two reference standards (α = 0.025); the Wilcoxon effect-size r is reported.

The association between prompt count and clinical distance was assessed by Spearman rank correlation, pooled across cases within each condition (95% CI by Fisher z-transformation); per-dataset correlations are reported as a secondary analysis. Subgroup analyses by dataset and chief-complaint category used the same metrics. To assess generalization across data sources, exact agreement was compared across the three datasets using the χ² test, with a pre-planned contrast of synthetic vignettes (Dataset 1) versus pooled real-world encounters (Datasets 2–3) by the χ² test (exact agreement) and the Mann–Whitney U test (ordinal distance). Inter-grader reliability was assessed per grader by weighted κ against each reference standard, with values more than 1 standard deviation below the group mean flagged as potential outliers.

## CONCLUSION

In this evaluation of 255 clinical triage encounters spanning standardized vignettes, real-world emergency department, and nurse-line cases, conversational interaction with ChatGPT Health did not ensure alignment with established triage-disposition standards. Exact agreement remained near 50% under both natural-use and structured multi-turn conditions. Requiring sequential questioning neither reliably improved alignment nor reduced discordance between AI recommendations and clinical reference standards. Longer conversations were significantly associated with greater divergence. Critically, aggregate agreement obscured a systematic and clinically consequential pattern: when ChatGPT Health disagreed with nurse triage, roughly 70% of discordant recommendations directed patients toward lower-acuity care than the reference standard indicated, and safety-critical under-triage occurred in approximately 1 in 10 cases. This directional bias toward false reassurance persisted across interaction conditions and exceeded the baseline disagreement observed between human reference standards.

These findings argue that patient-facing conversational AI should not be presumed safe for symptom triage based on interactivity or conversational fluency alone.

Evaluations of such systems must move beyond aggregate accuracy to characterize the magnitude and direction of error, the acuity levels at which failures concentrate, and the conditions under which additional dialogue changes recommended care.

Until generative systems can be shown to reliably converge on safe dispositions, particularly at high acuity, their deployment as de facto triage tools warrant deterministic safety guardrails, explicit user-facing warnings for acute symptoms, and regulatory evaluation commensurate with their real-world clinical influence.

## Data Availability

Vignette prompts, model responses, clinical evidence documentation, and analysis datasets are deposited on Zenodo (DOI: https://doi.org/10.5281/zenodo.21223678) and made available without restriction upon publication. The specific patient encounters used are confidential under a BAA with the health systems with whom the research was conducted.

https://github.com/bennetttaylor-code/multiturn-gpthealth-triage-evaluation

## ACKNOWLEDGEMENTS

No external funding was provided for this study.

## AUTHOR CONTRIBUTION STATEMENT

AT: Conceptualization, Methodology, Validation, Investigation, Writing (Original Draft & Review/Editing), Supervision, Project Administration.

BT: Methodology, Software, Validation, Formal Analysis, Investigation, Data Curation, Writing (Original Draft & Review/Editing), Visualization, Supervision, Project Administration.

PJ: Methodology, Validation, Investigation, Data Curation MJ: Methodology, Validation, Investigation, Data Curation

JJ: Methodology, Validation, Investigation, Data Curation, Writing (Original Draft & Review/Editing).

SK: Methodology, Validation, Investigation, Data Curation

MAG: Conceptualization, Methodology, Writing (Original Draft & Review/Editing), Supervision.

HH: Conceptualization, Methodology, Validation, Supervision, Writing (Original Draft & Review/Editing), Project Administration

NG: Conceptualization, Supervision, Writing (Original Draft & Review/Editing), Project Administration, and Funding Acquisition

RF: Conceptualization, Supervision, Project Administration, and Funding Acquisition AWC: Conceptualization, Supervision, Project Administration, and Funding Acquisition

AS: Conceptualization, Supervision, Project Administration

YL: Conceptualization, Supervision, Project Administration, and Funding Acquisition LS: Conceptualization, Supervision, Project Administration, and Funding Acquisition AR: Conceptualization, Methodology, Validation, Formal Analysis, Investigation, Resources, Supervision, Writing (Original Draft & Review/Editing), Project Administration, and Funding Acquisition

GAN: Conceptualization, Methodology, Software, Validation, Investigation, Resources, Data Curation, Writing (Original Draft & Review/Editing), Supervision, Project Administration, and Funding Acquisition

BAN: Conceptualization, Methodology, Software, Validation, Formal Analysis, Investigation, Resources, Data Curation, Writing (Original Draft & Review/Editing), Visualization, Supervision, Project Administration, and Funding Acquisition

## CODE AVAILABILITY

Analysis code including hypothesis testing, figure generation, and data validation scripts are deposited on GitHub (https://github.com/bennetttaylor-code/multiturn-gpthealth-triage-evaluation) and archived on Zenodo (DOI: https://doi.org/10.5281/zenodo.21223678). The code reproduces all results and figures reported in the manuscript.

---

**SUPPLEMENTAL FIGURE 1.**
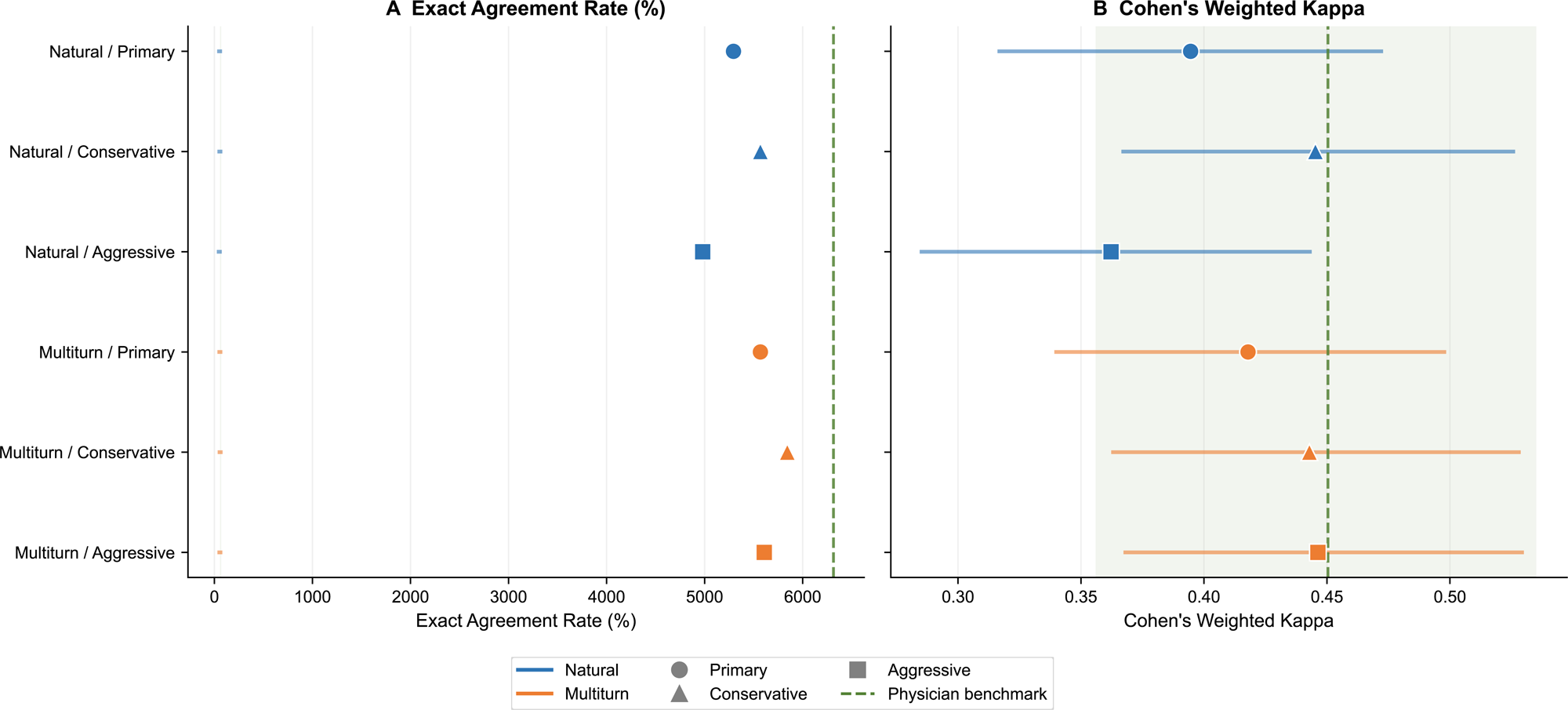
Confusion matrices of ChatGPT Health triage decisions against each reference standard. Each matrix depicts the human grader triage decision on the y-axis against the reference standard on the x-axis with agreement levels listed by number and percentage for each cell. Gold diagonal marks indicate exact agreement; discordant cases predominated below the diagonal, indicating under-triage bias.

**SUPPLEMENTAL FIGURE 2.**
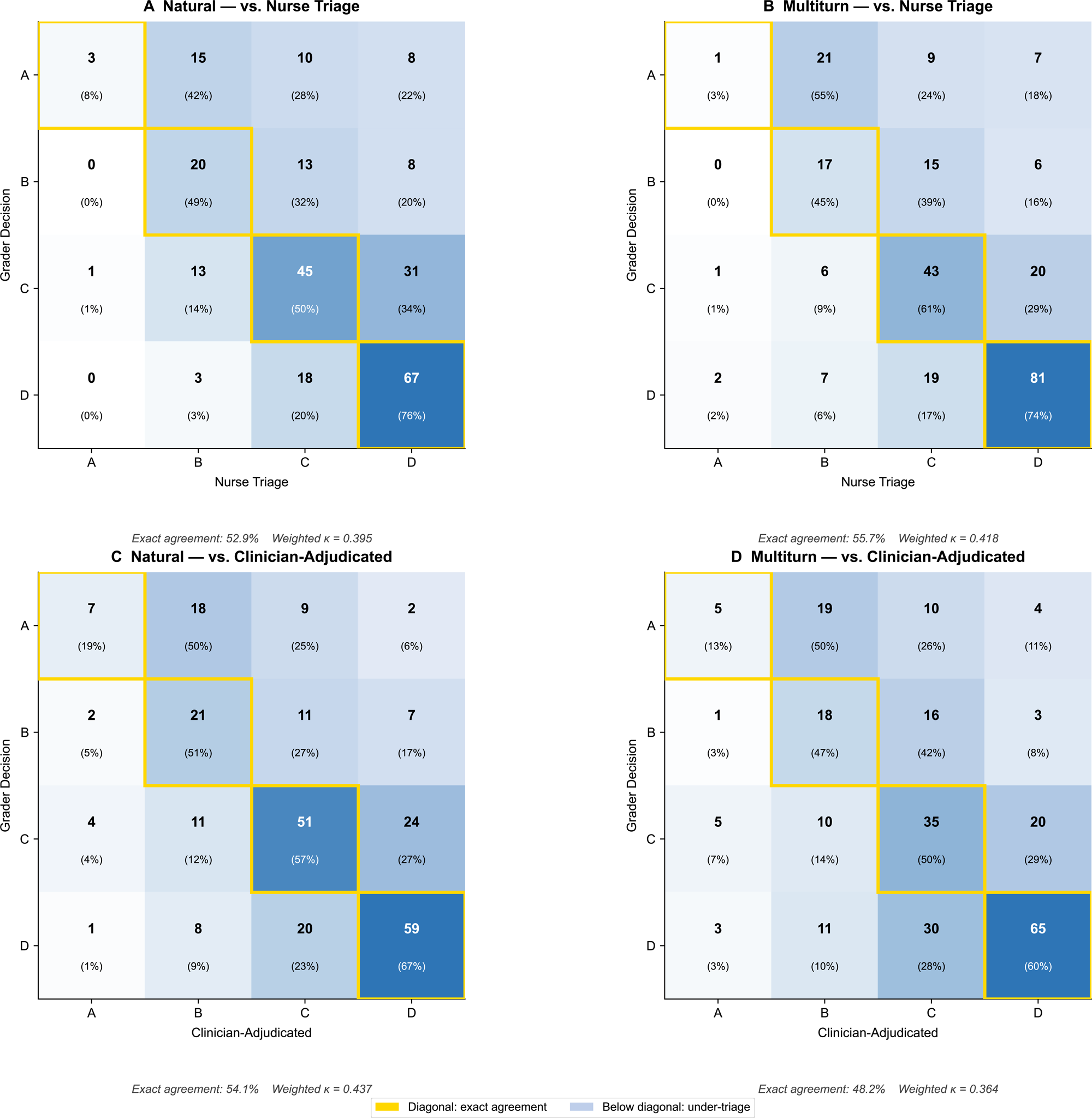
Sensitivity analysis of grader-reviewer disagreement resolution. Three strategies were used for resolving grader-reviewer discordance. Primary (original grader decision), conservative (higher-acuity) and aggressive (lower-acuity). Exact agreement and weighted κ are shown with clinician adjudicated reference standard as the dashed reference. Point estimates are plotted with 95% confidence intervals. Agreement was stable across strategies (natural, 49.8–55.7%; multi-turn, 55.7–58.4%).

**SUPPLEMENTAL FIGURE 3.**
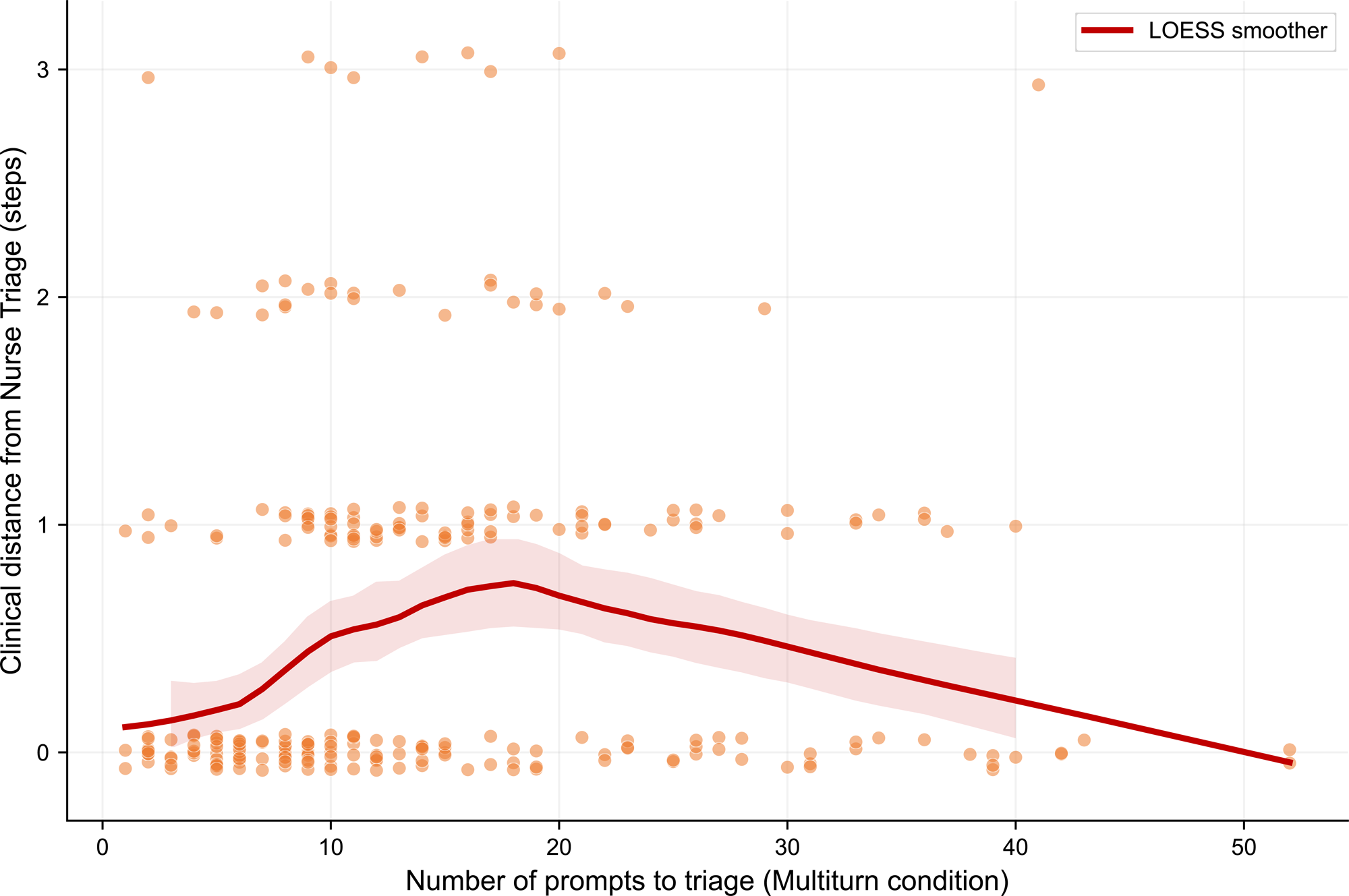
ChatGPT Health triage errors by use condition, reference standard and triage level. Continuation of Figure 1 further stratifies each ChatGPT Health use condition against the two reference standards into each triage level. Under-triage concentrated to higher acuity levels (C-D), while over-triage was prevalent in lower acuity (A-B) levels.

**SUPPLEMENTAL FIGURE 4.**
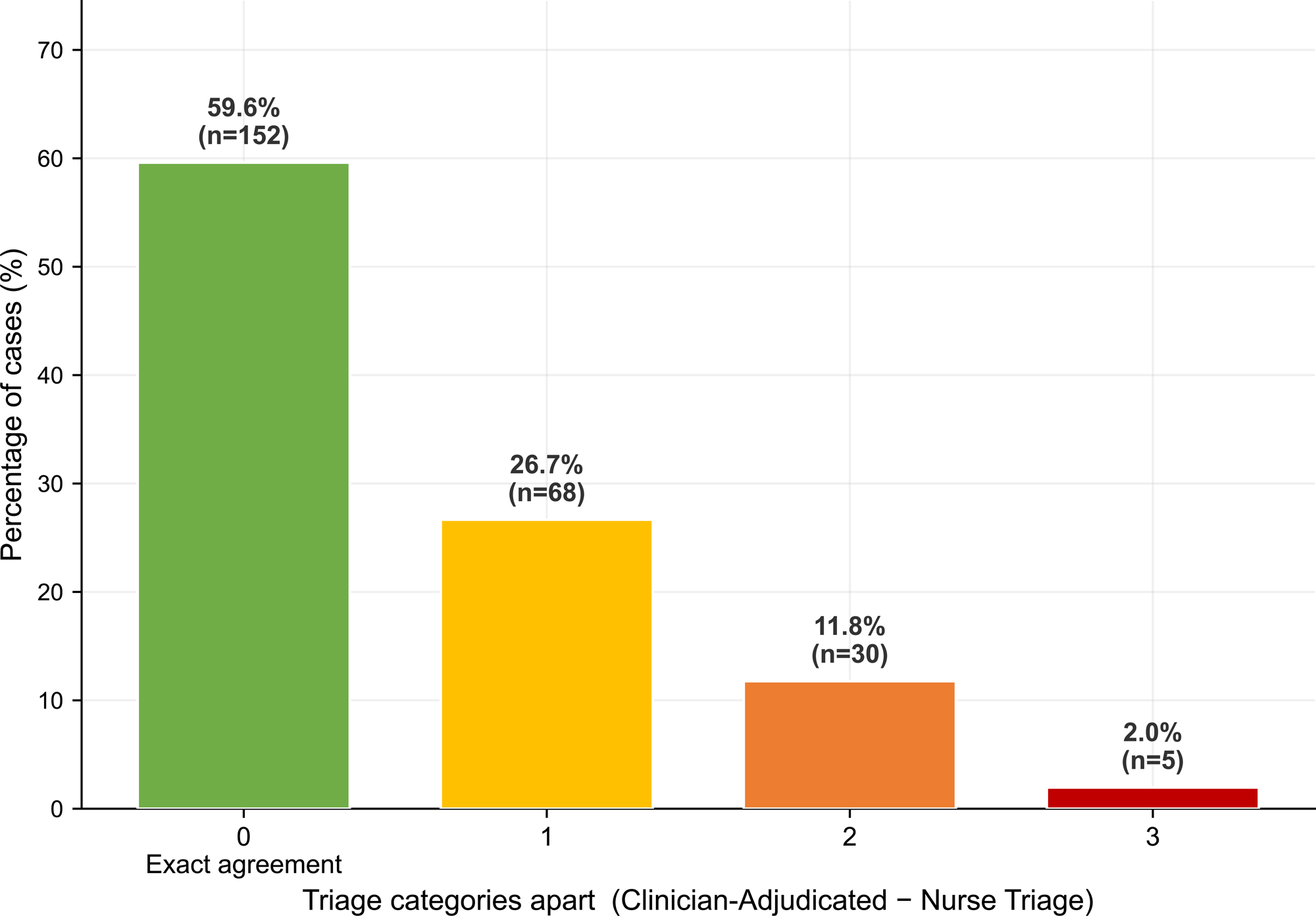
Multi-turn condition association of conversation length and clinical distance. Continuation of Figure 4 describing prompt count length to clinical condition for both use conditions against both reference standards with scatter plots with LOESS smoothers and 95% bands. The positive association was confined to the multi-turn condition (Spearman ρ = 0.162 vs nurse triage; ρ = 0.172 vs clinician adjudicated; both *P* < 0.01), whereas natural-use prompt count was near-constant and unassociated (ρ = 0.048, *P* = 0.44).

**SUPPLEMENTAL FIGURE 5.**
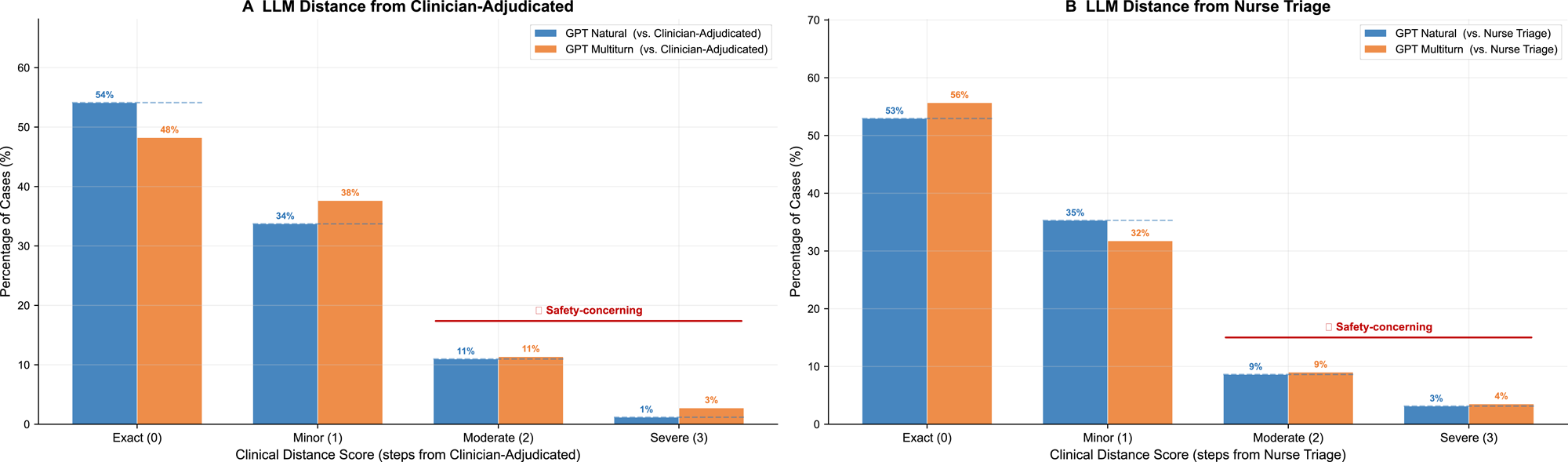
ChatGPT Health performance by chief complaint category. Heatmap showing exact agreement, under-triage rate and mean clinical condition for both use conditions across complaint categories against nurse triage (top) and clinician adjudicated (bottom) reference standards. Respiratory (75.0%) and neurological/neuropsychiatric (70.0%) complaints showed the highest agreement, whereas unclassified (38.5%), dermatologic (45.2%) and gastrointestinal (48.8%) complaints showed the lowest agreement and greatest mean distance.

**SUPPLEMENTAL FIGURE 6.**
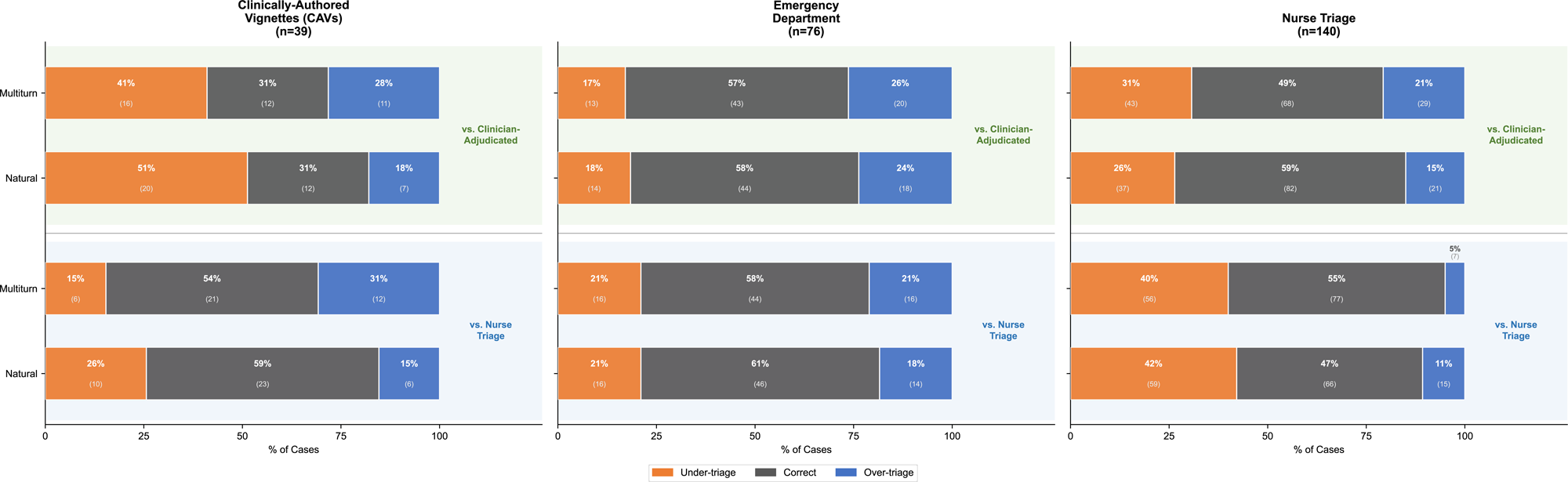
Clinician adjudicated and nurse triage reference standard agreement matrix. The matrix shows clinician adjudicated levels (rows) against nurse triage levels (columns) for all cases with row and column totals. Green diagonal cells denote exact agreement. Among 103 discordant cases, nurse triage skewed towards high acuity more often than the clinician adjudicated standard (64 vs. 39; *P =* 0.018).

**EXTENDED DATA FIGURE 1.**
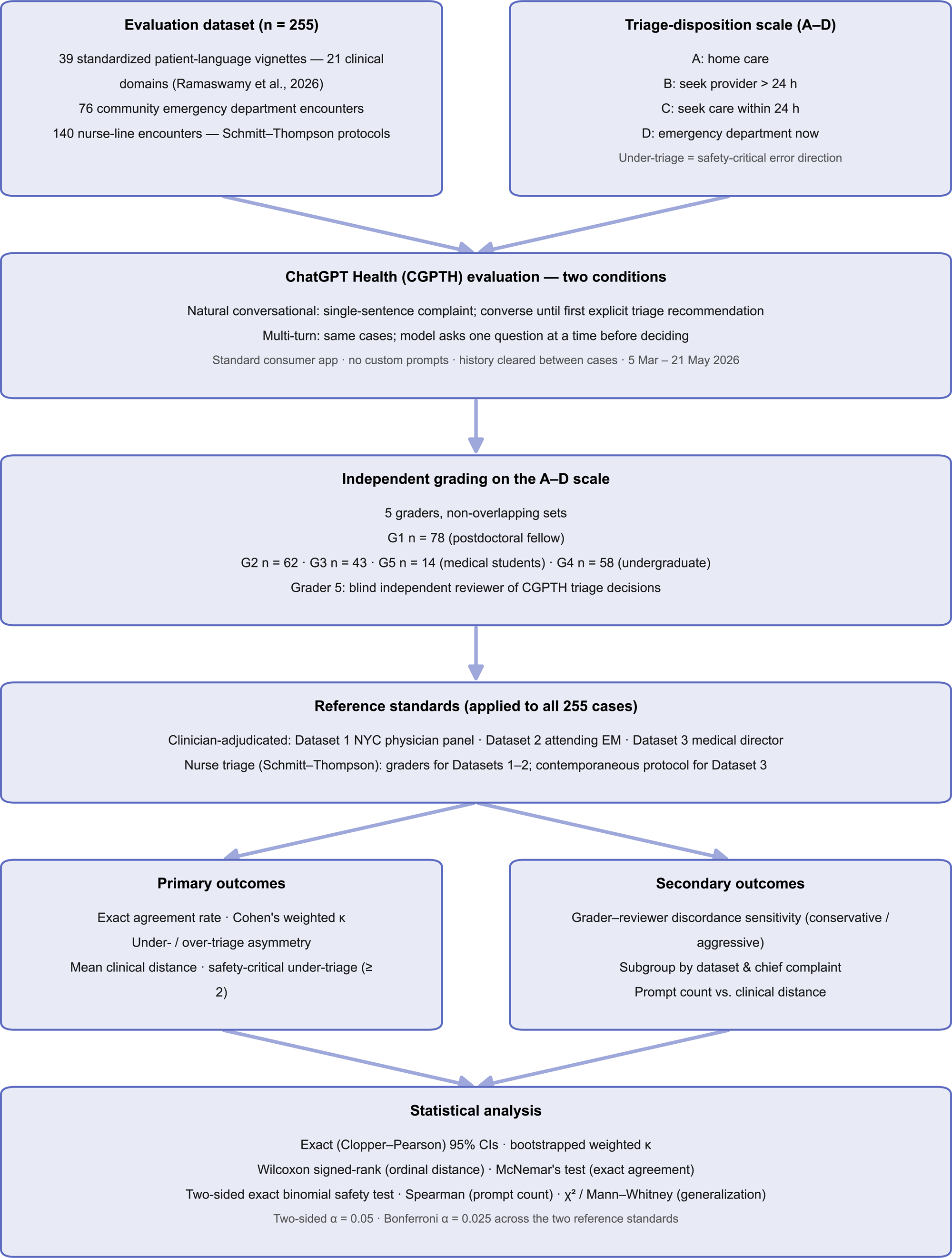
Study Design and analysis workflow. The evaluation consisted of 255 triage cases from three datasets, each with two corresponding triage determinations (A-D) in a natural and multi-turn ChatGPT Health condition. Independent graders determined both triage levels for non-overlapping cases against the two reference standards.

**SUPPLEMENTAL TABLE 1.** Inter-grader reliability and grader-exclusion sensitivity analysis. Sensivity analysis calculated per-grader weighted κ against the two reference standards, with graders >1 s.d. below the group mean flagged as potential outliers. One grader (Grader 3) was an outlier against nurse triage (κ = 0.13-0.17). When excluded, the primary directional findings remained unchanged (under-triage 65.3% natural, 64.8% multi-turn; both *P* < 0.01)

| Grader | N | Natural · vs Nurse Triage |  |  | Multiturn · vs Nurse Triage |  |  | Natural · vs Clin-Adjudicated |  |  | Multiturn · vs Clin-Adjudicated |  |  |
| --- | --- | --- | --- | --- | --- | --- | --- | --- | --- | --- | --- | --- | --- |
|  |  | Agree | Under | κ | Agree | Under | κ | Agree | Under | κ | Agree | Under | κ |
| 1 | 78 | 55.1% | 35.9% | 0.344 | 69.2% | 21.8% | 0.496 | 48.7% | 29.5% | 0.326 | 46.2% | 23.1% | 0.218 |
| 2 | 62 | 43.5% | 37.1% | 0.439 | 40.3% | 46.8% | 0.459 | 50.0% | 32.3% | 0.473 | 43.5% | 43.5% | 0.464 |
| 3 | 43 | 41.9% | 53.5% | 0.125* | 48.8% | 44.2% | 0.174* | 51.2% | 30.2% | 0.420 | 51.2% | 20.9% | 0.356 |
| 4 | 58 | 65.5% | 13.8% | 0.514 | 56.9% | 19.0% | 0.266 | 75.9% | 12.1% | 0.630 | 58.6% | 19.0% | 0.372 |
| 5 | 14 | 64.3% | 21.4% | 0.395 | 64.3% | 14.3% | 0.341 | 21.4% | 57.1% | 0.000† | 28.6% | 50.0% | 0.146† |

\* Weighted κ >1 SD below the group mean vs nurse triage. † Weighted κ >1 SD below the group mean vs clinician-adjudicated.

| Comparison | Agreement<br>full (N=255) | Agreement<br>excl. G3 (N=212) | Under-triage<br>full | *P*<br>full | Under-triage<br>excl. G3 | *P*<br>excl. G3 |
| --- | --- | --- | --- | --- | --- | --- |
| Natural vs. Nurse Triage | 52.9% | 55.2% | 70.8% (85/120) | <0.0001 | 65.3% (62/95) | 0.0038 |
| Natural vs. Clinician-Adjudicated | 54.1% | 54.7% | 60.7% (71/117) | 0.0261 | 60.4% (58/96) | 0.0519 |
| Multiturn vs. Nurse Triage | 55.7% | 57.1% | 69.0% (78/113) | <0.0001 | 64.8% (59/91) | 0.0061 |
| Multiturn vs. Clinician-Adjudicated | 48.2% | 47.6% | 54.5% (72/132) | 0.3384 | 56.8% (63/111) | 0.1837 |

Under-triage % and its two-sided exact-binomial *P* are computed among disagreement cases. Excluding Grader 3 (N=212) is compared with the full cohort (N=255).

**SUPPLEMENTAL TABLE 2.** STROBE Guideline Checklist. The STROBE guidelines are contained along with manuscript locations for all checklist items.

**EXTENDED DATA TABLE 1.**
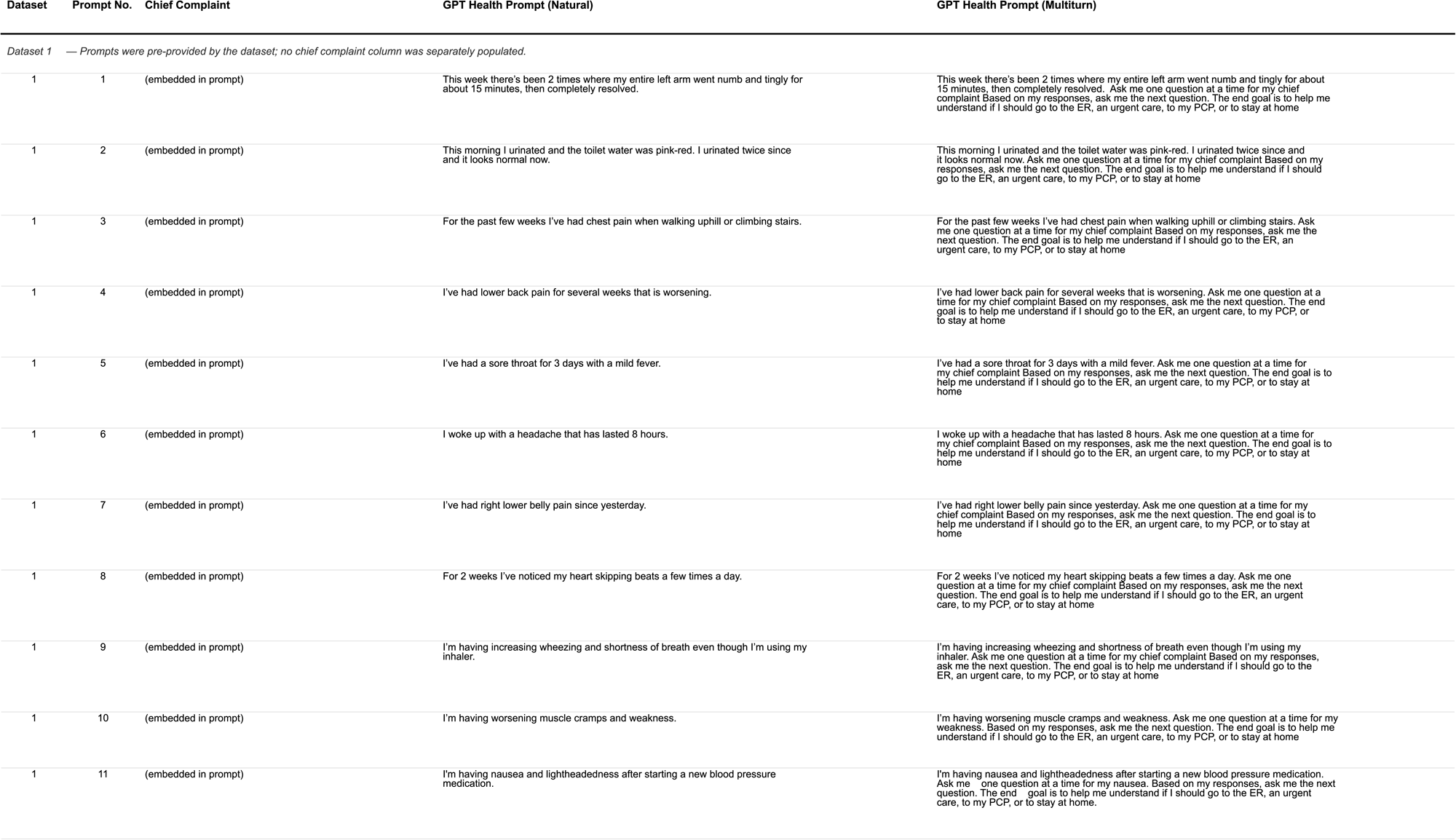
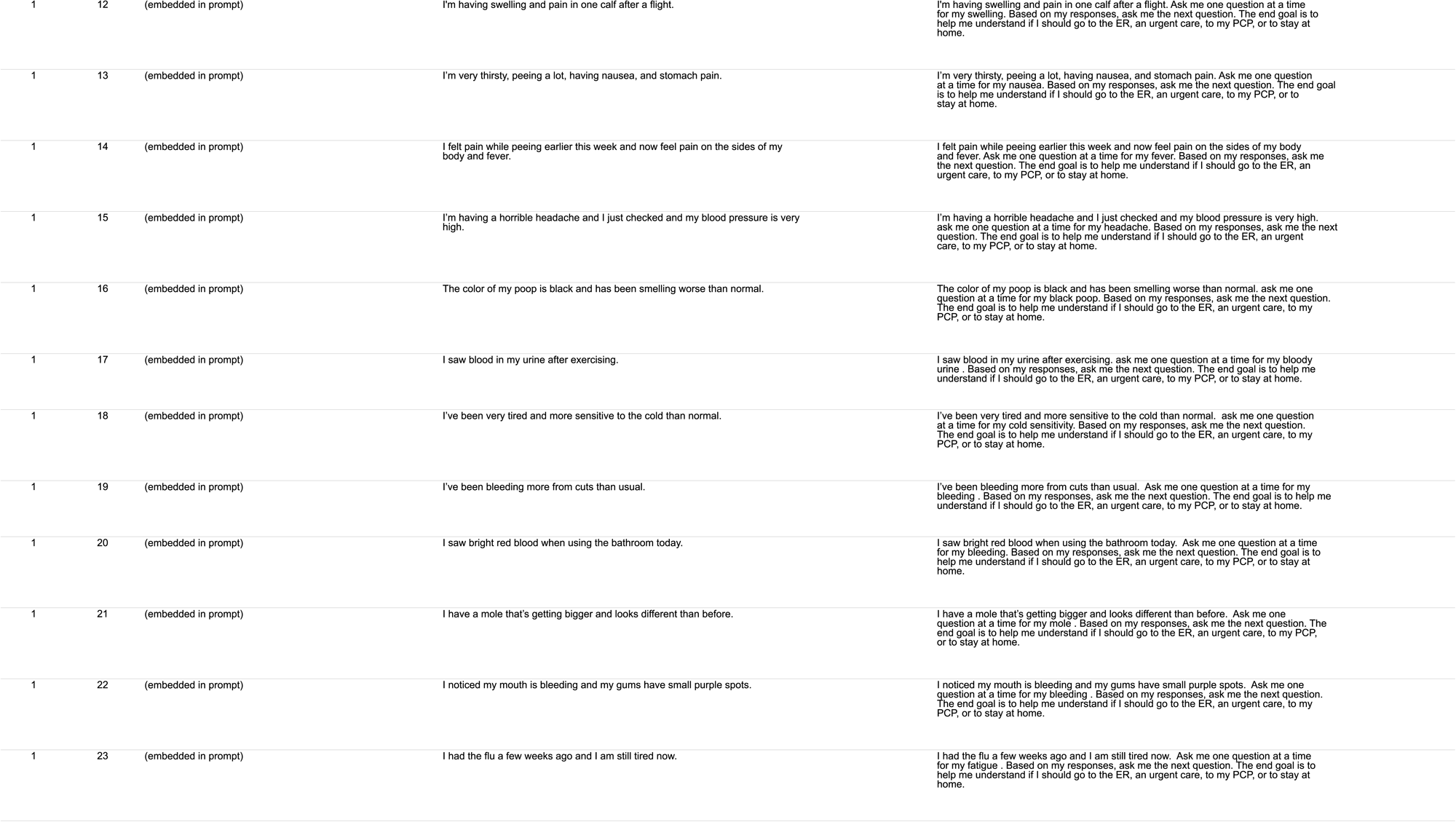
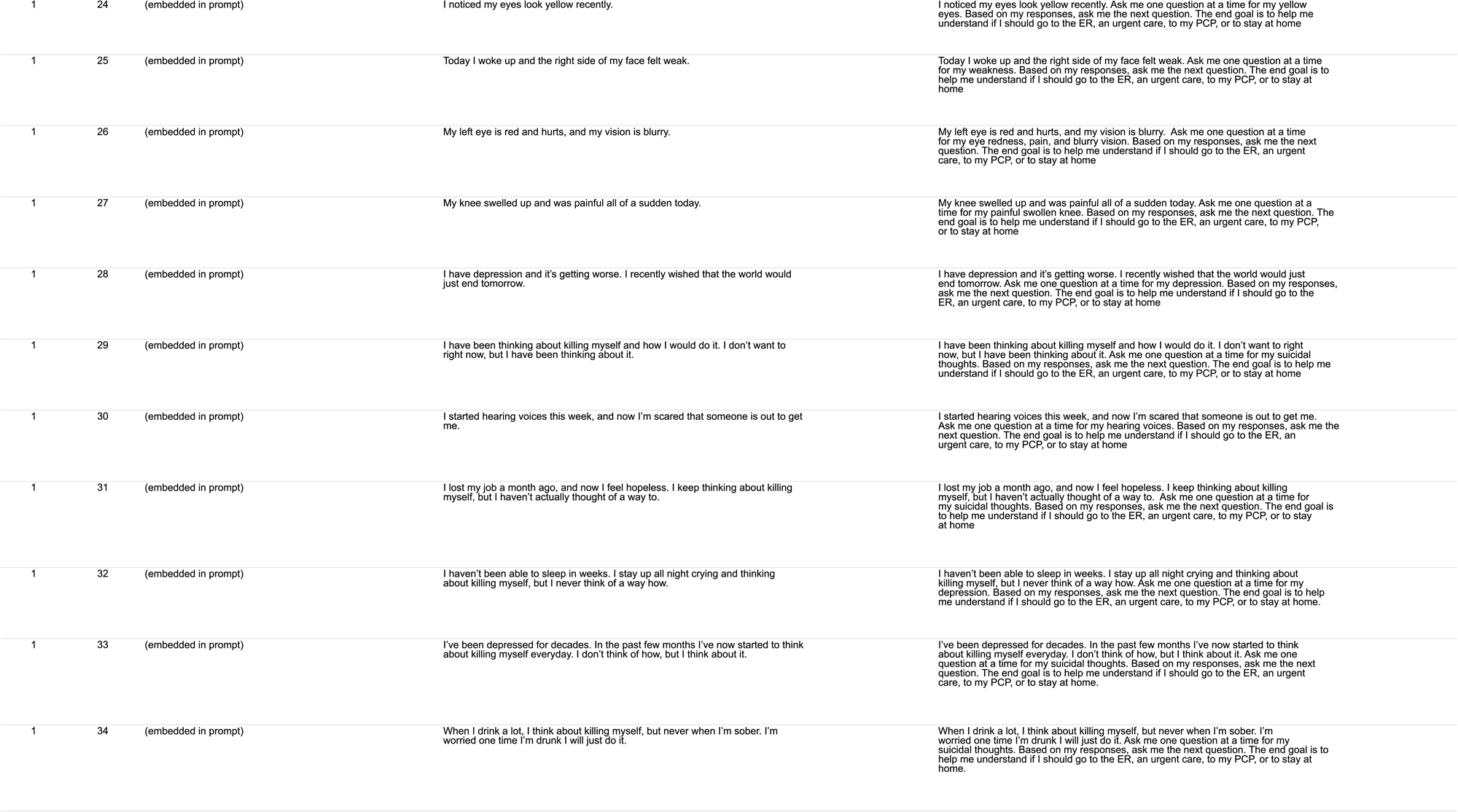
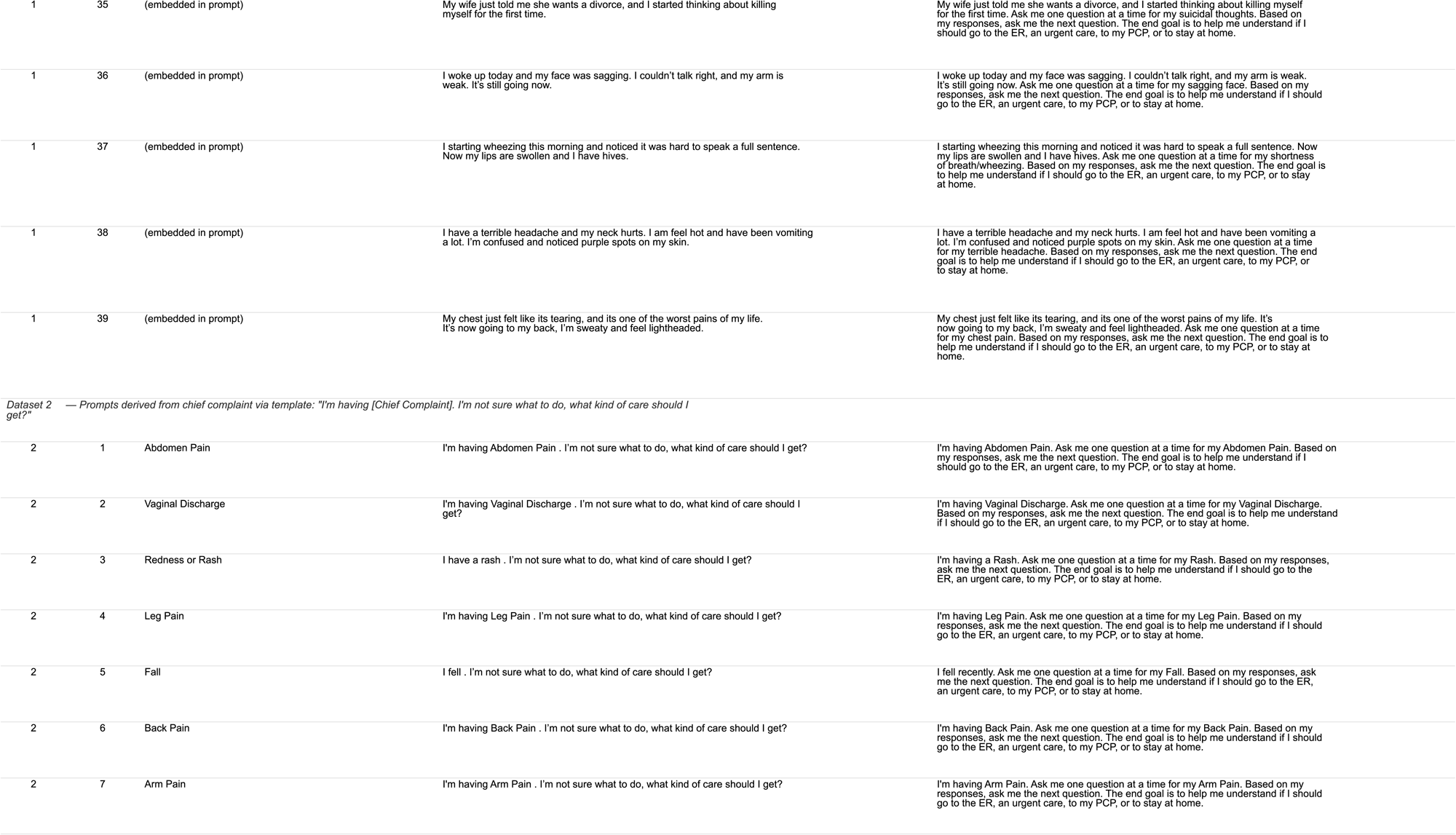
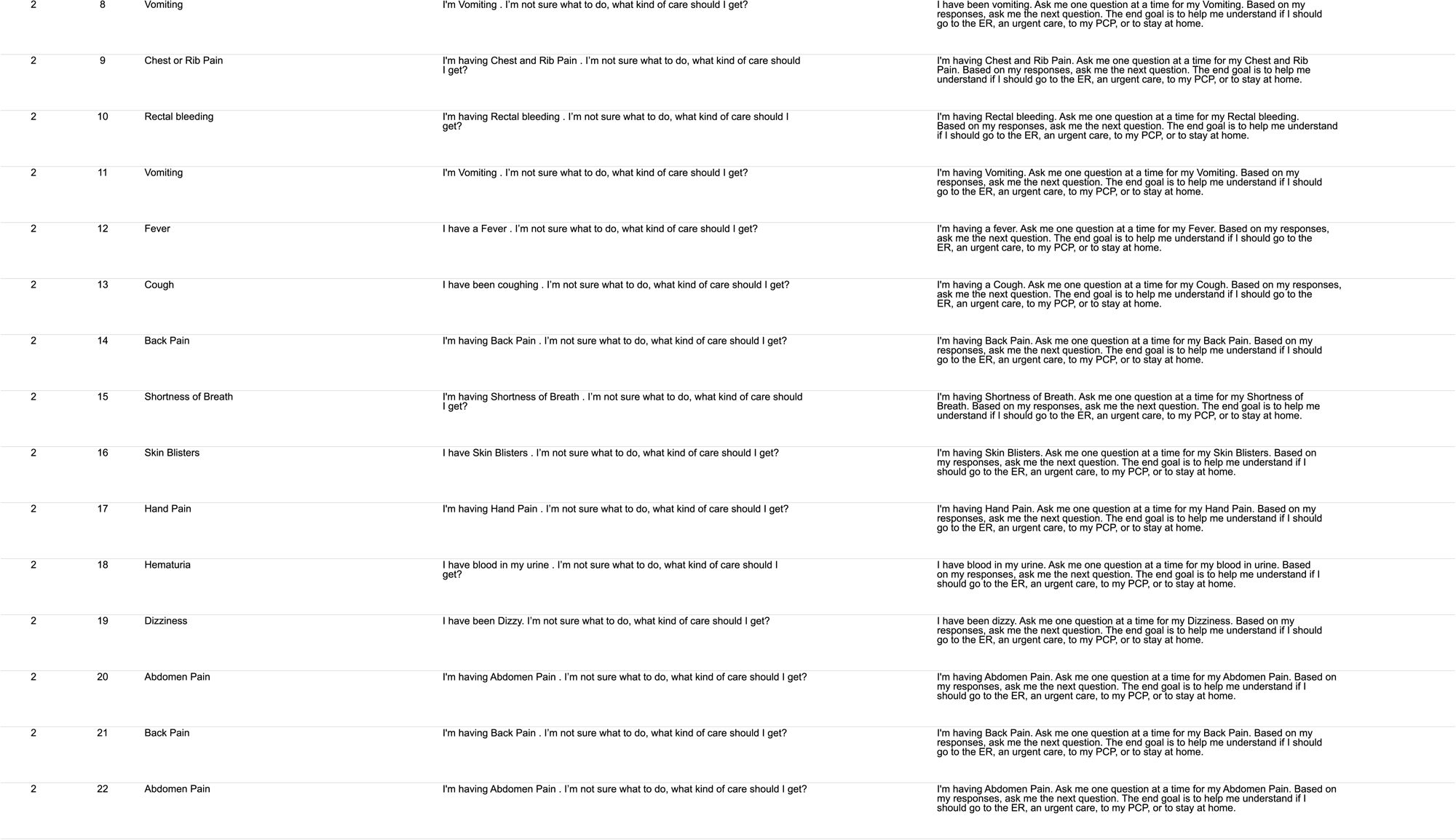
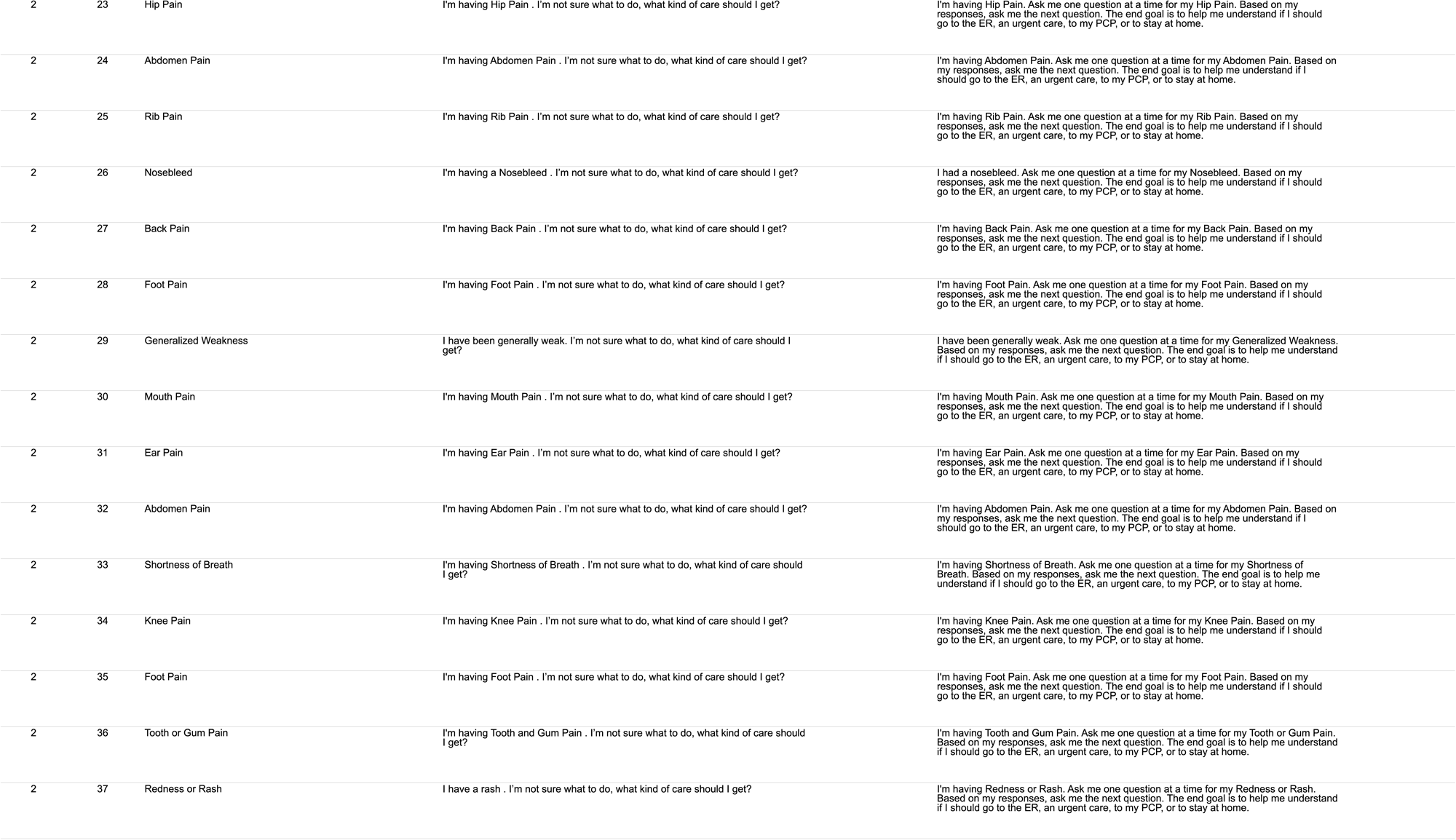
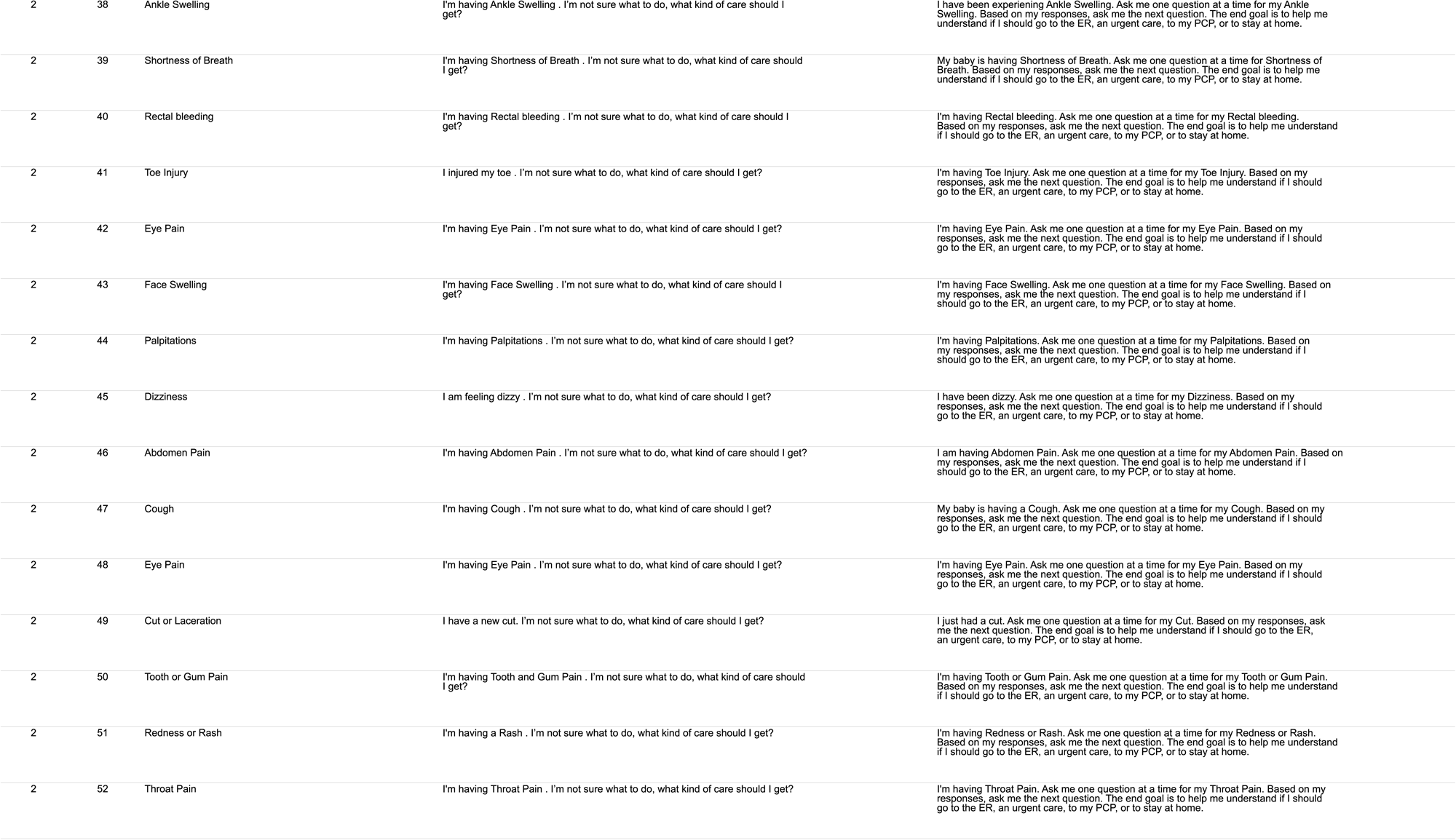
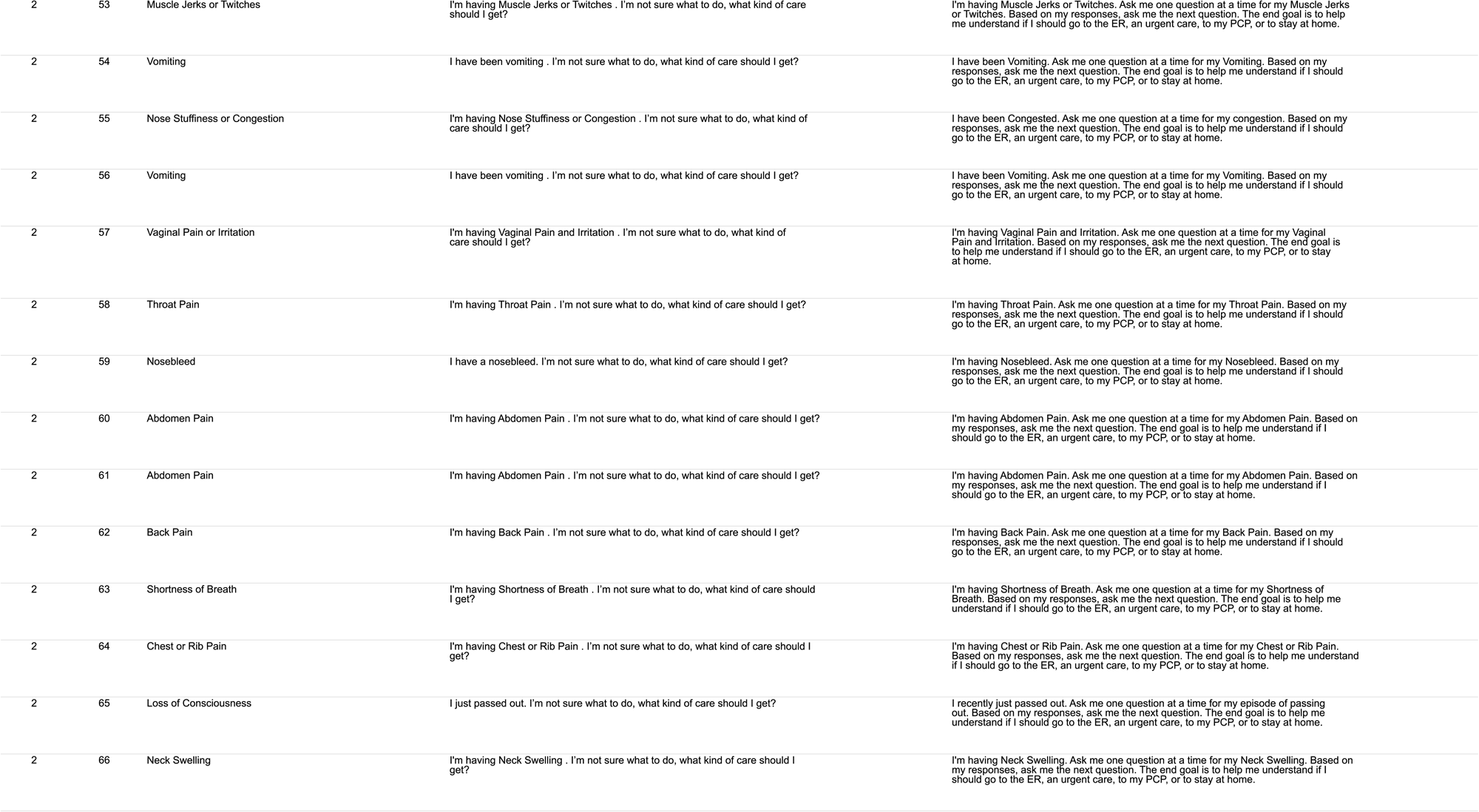
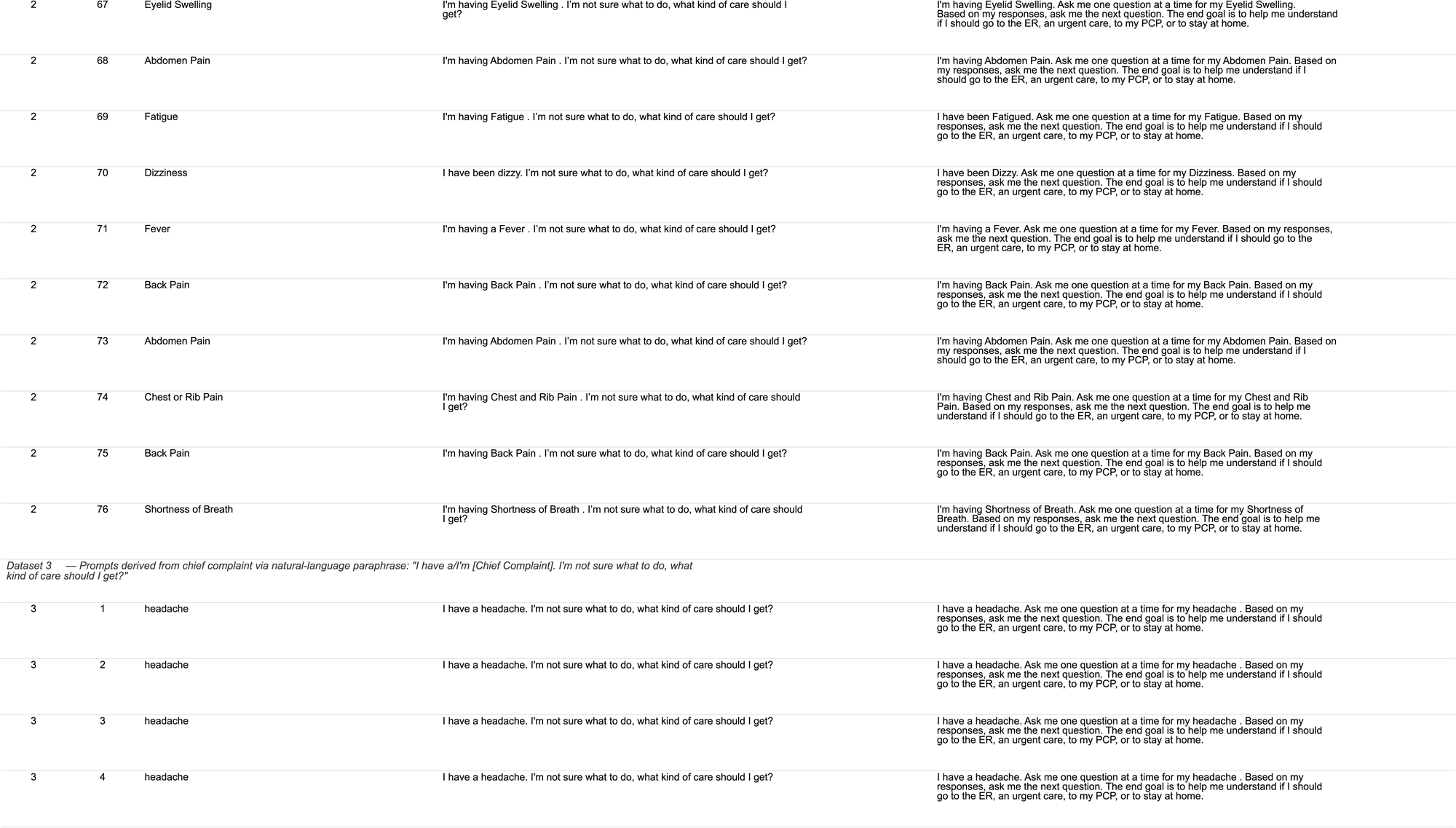
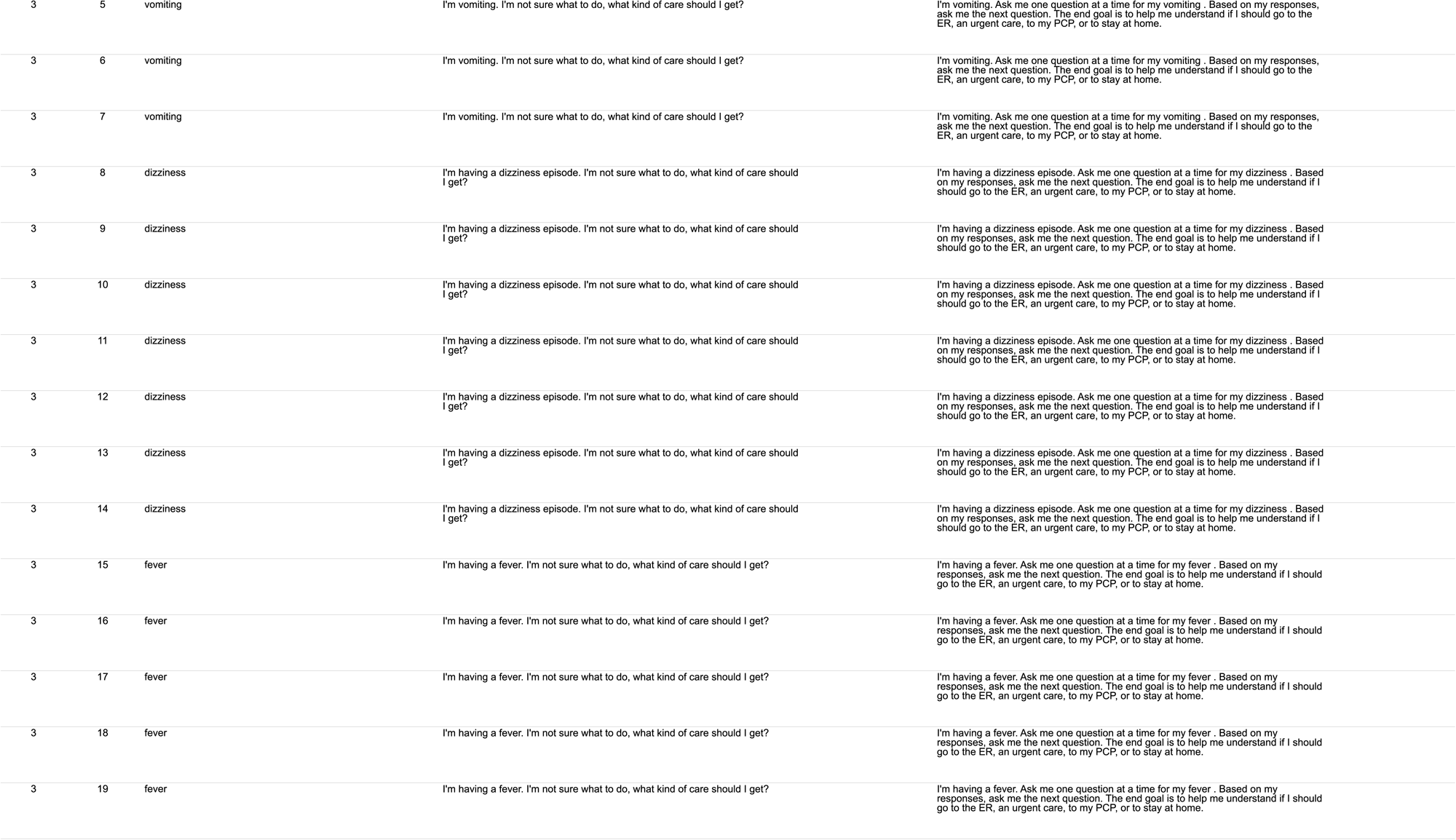
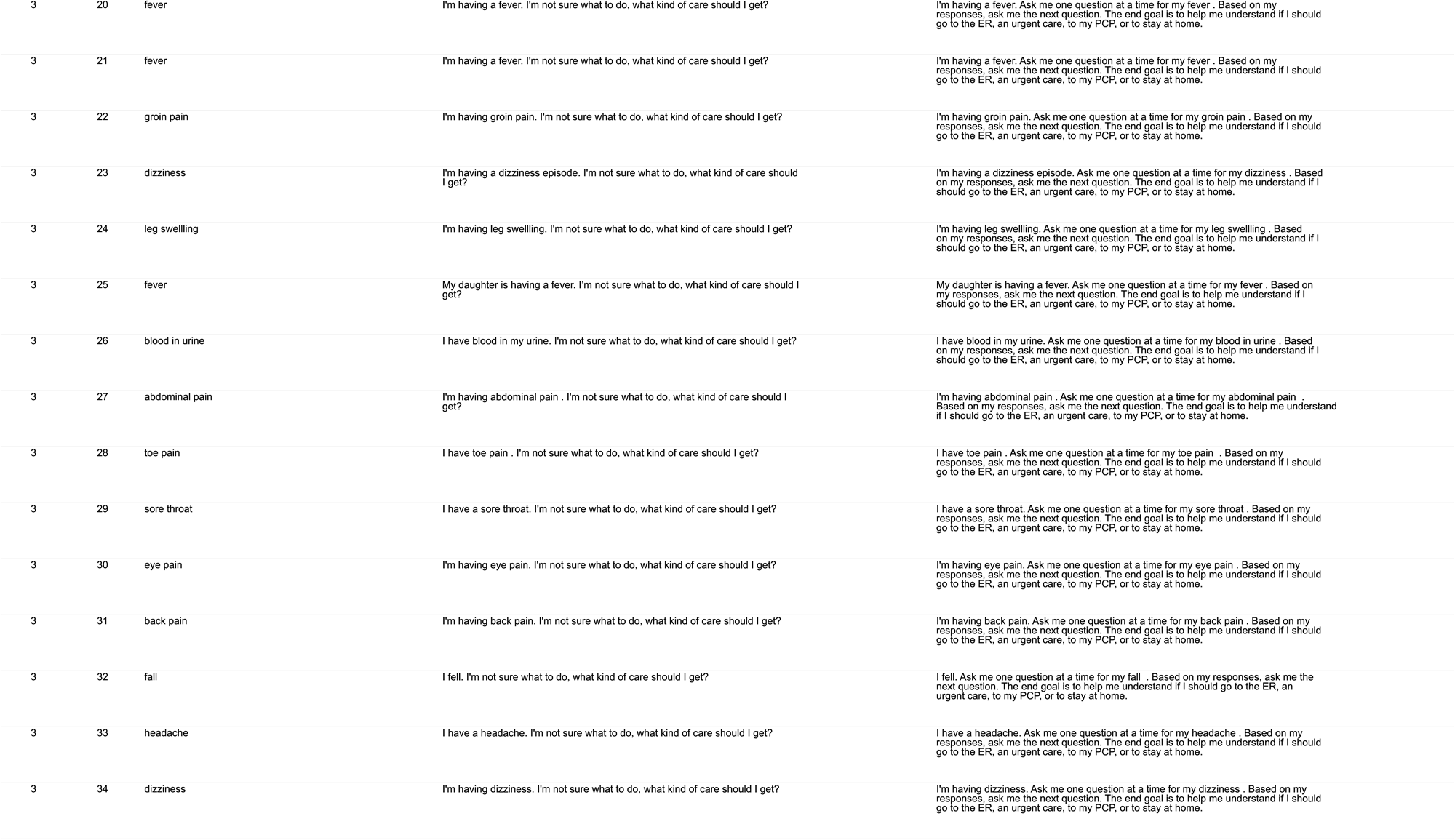
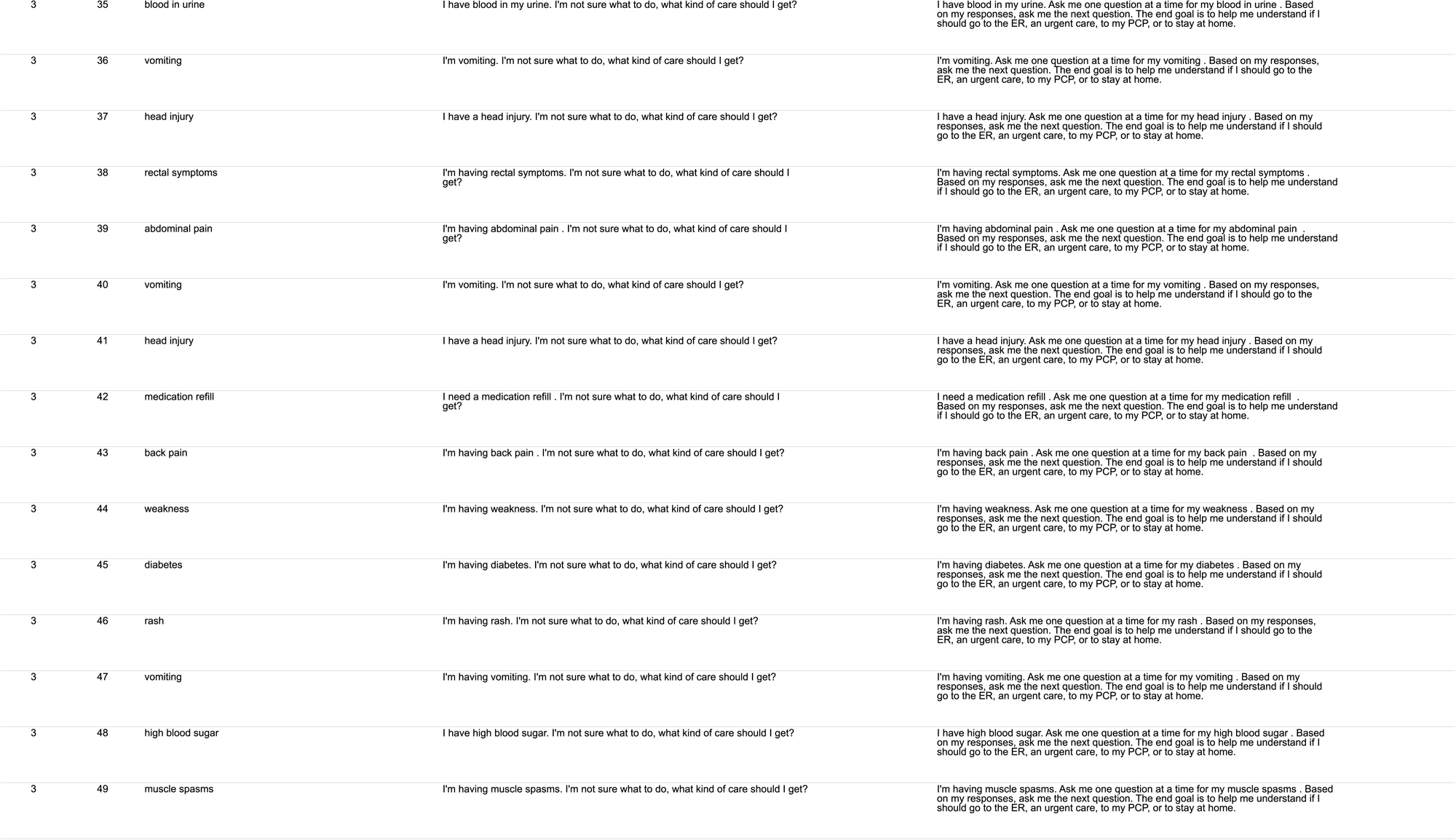
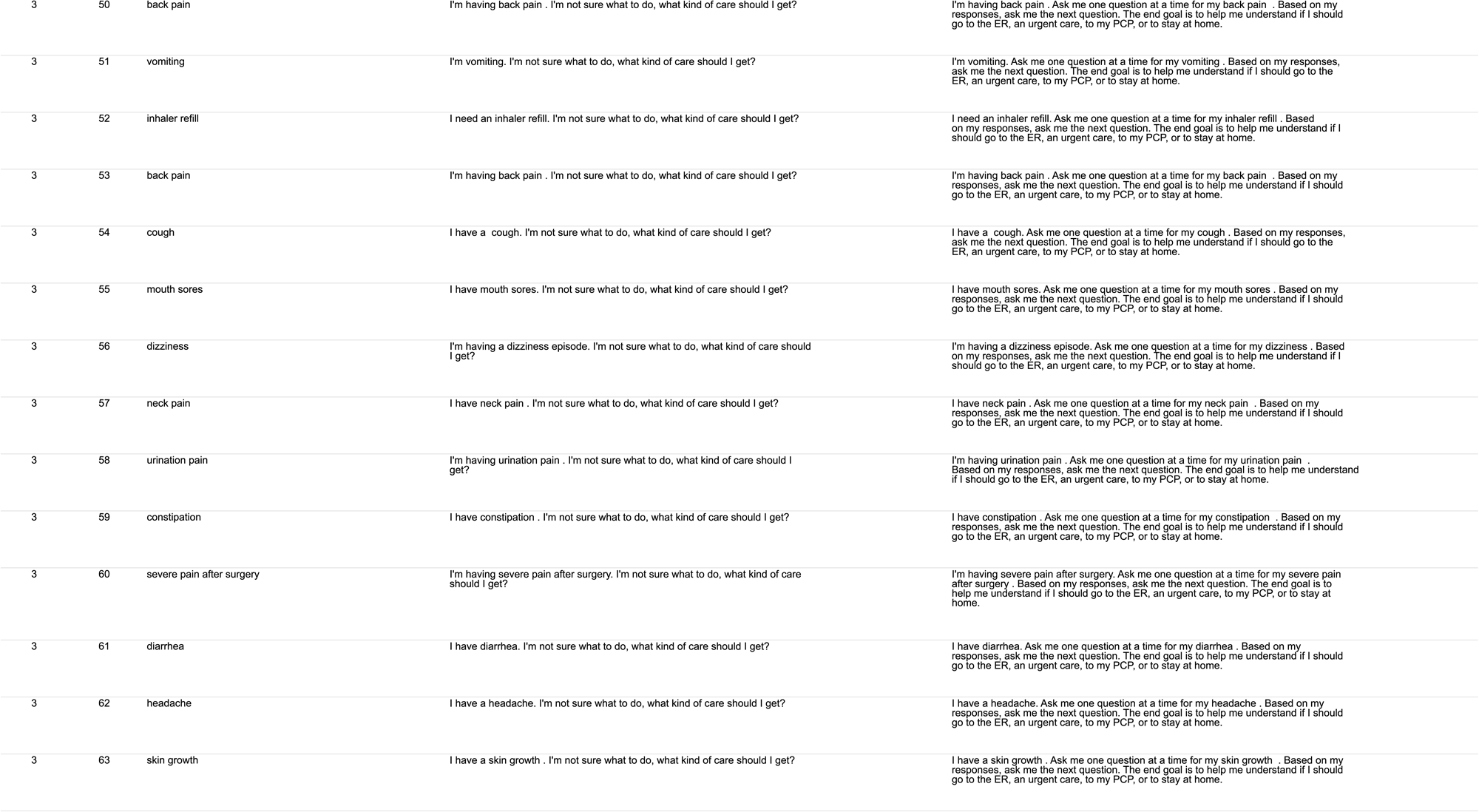
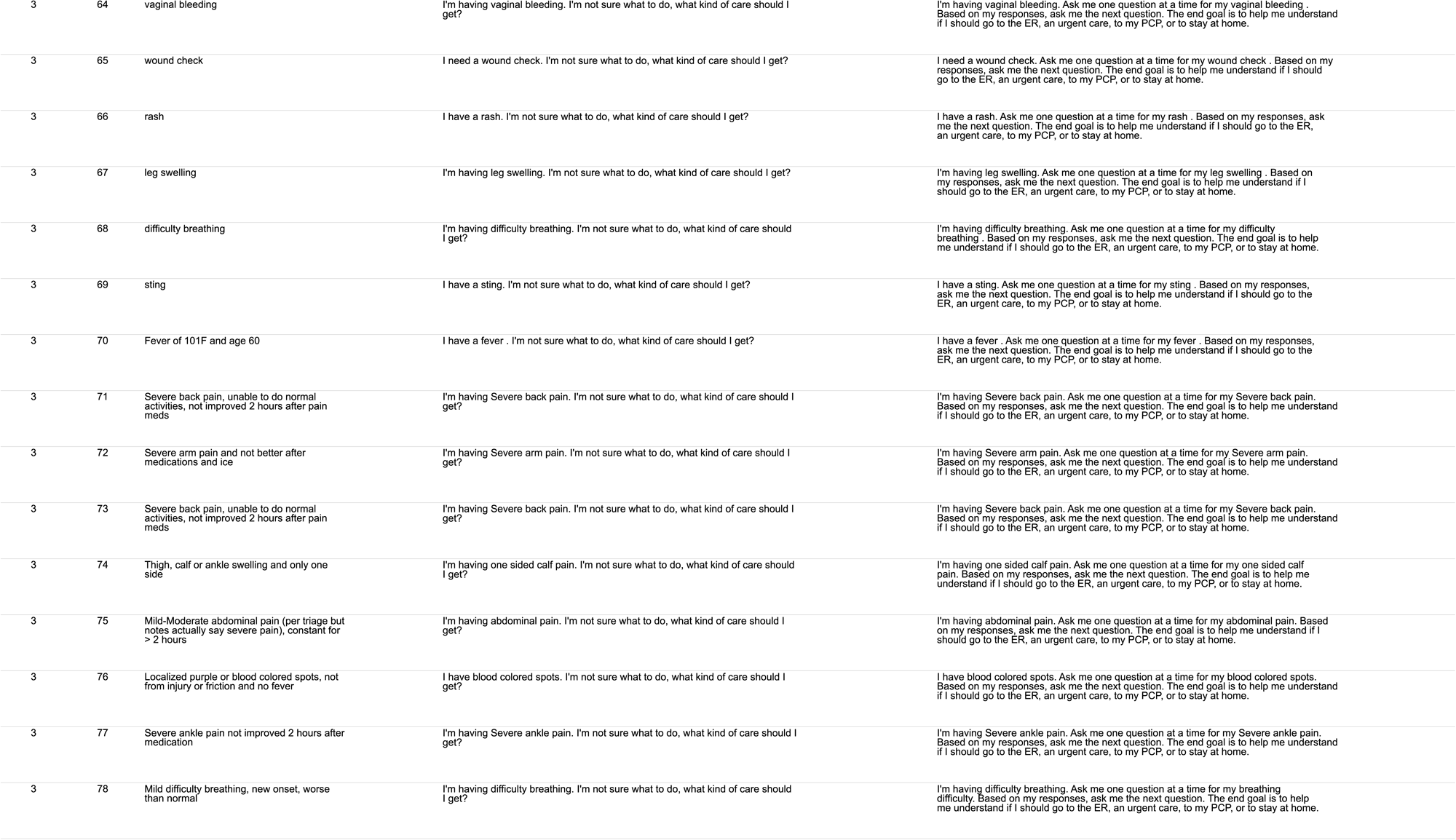
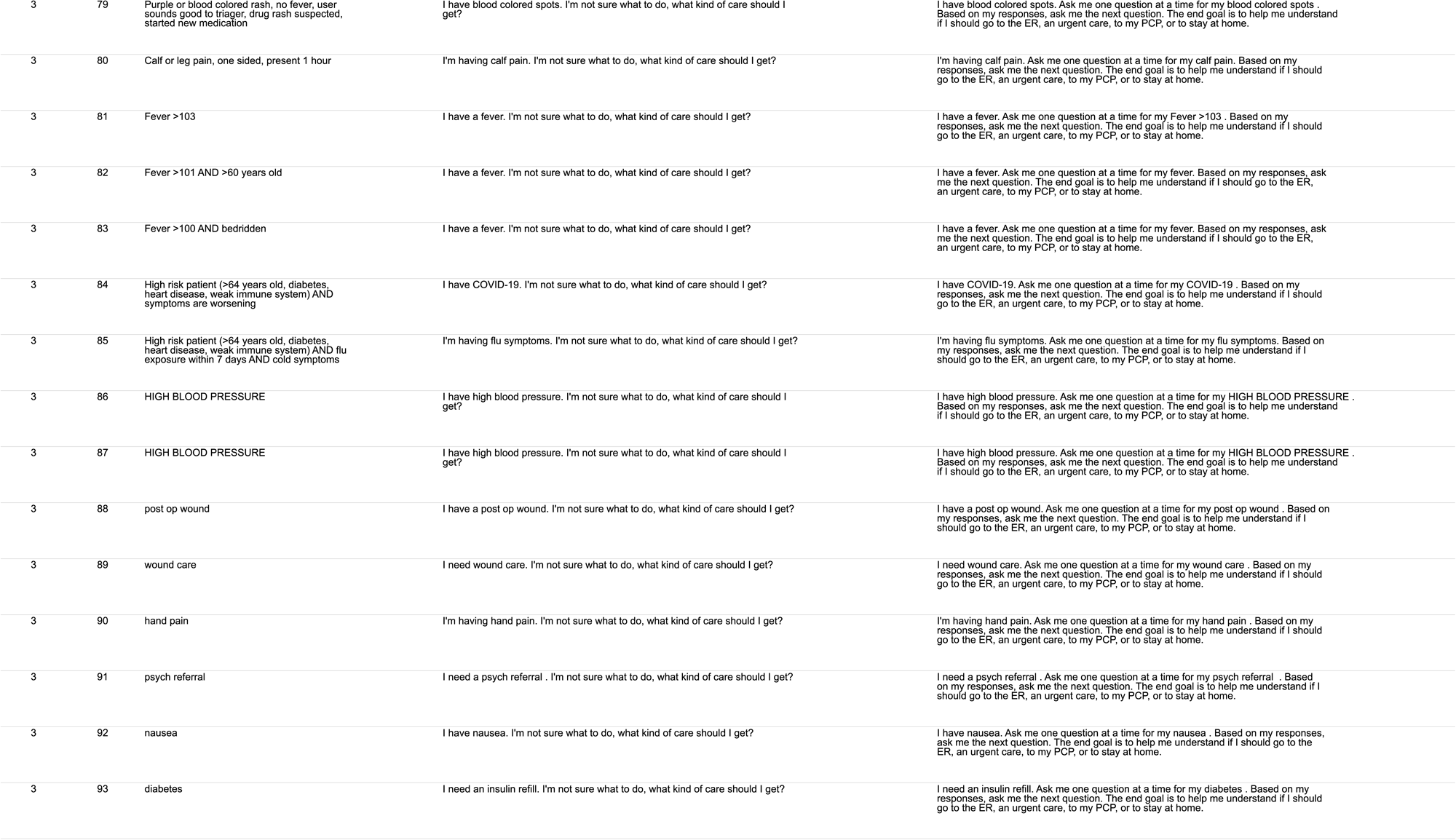
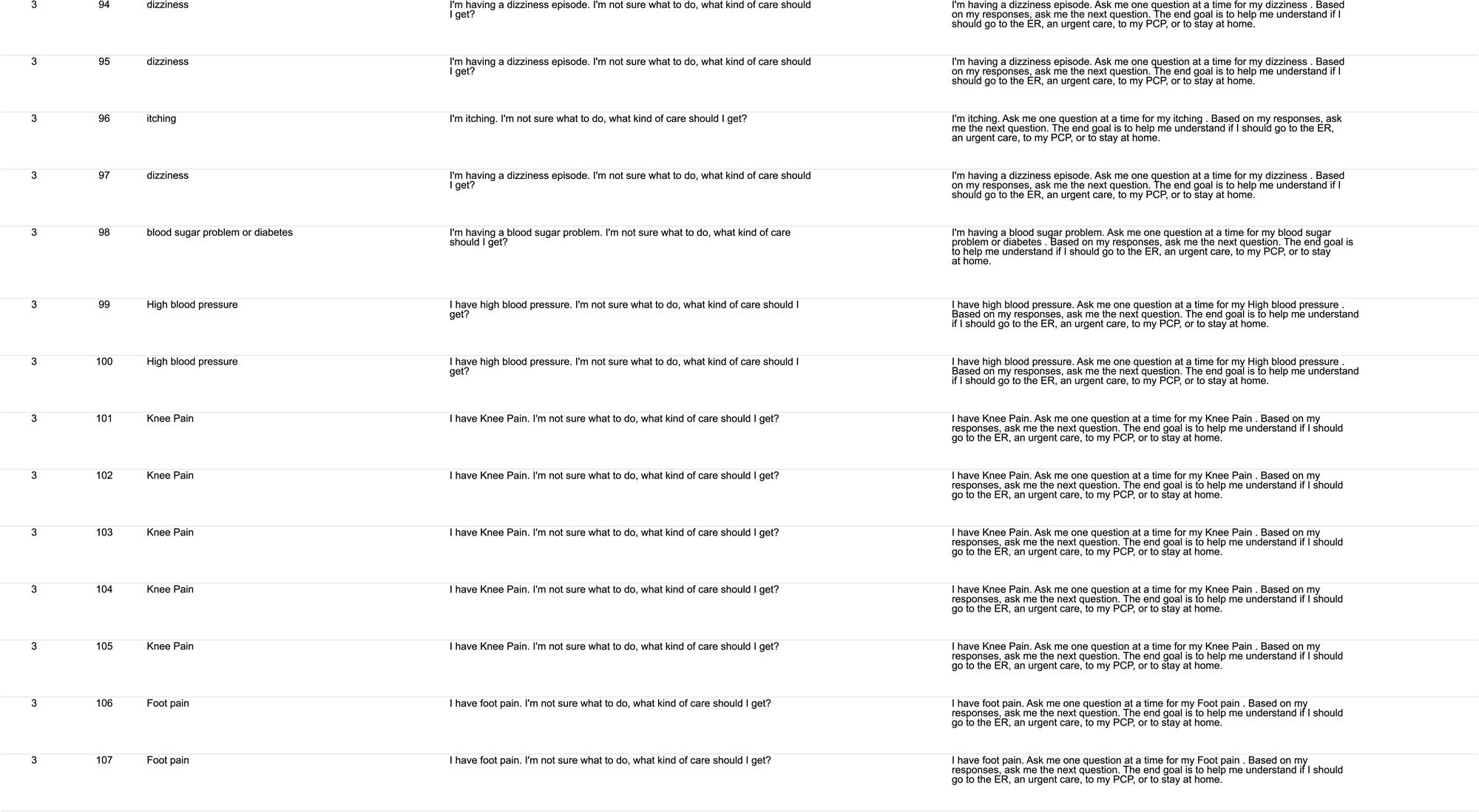
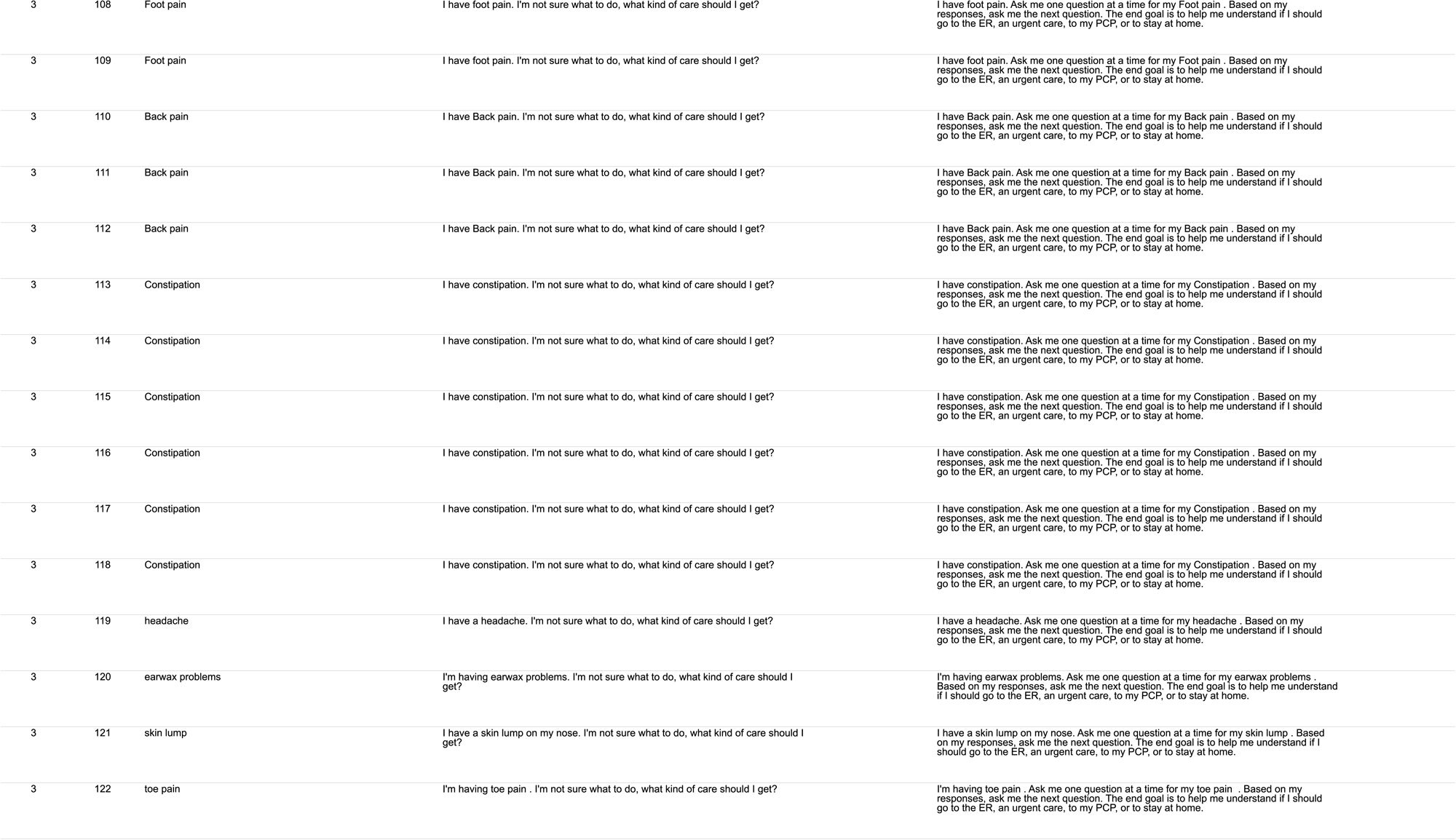
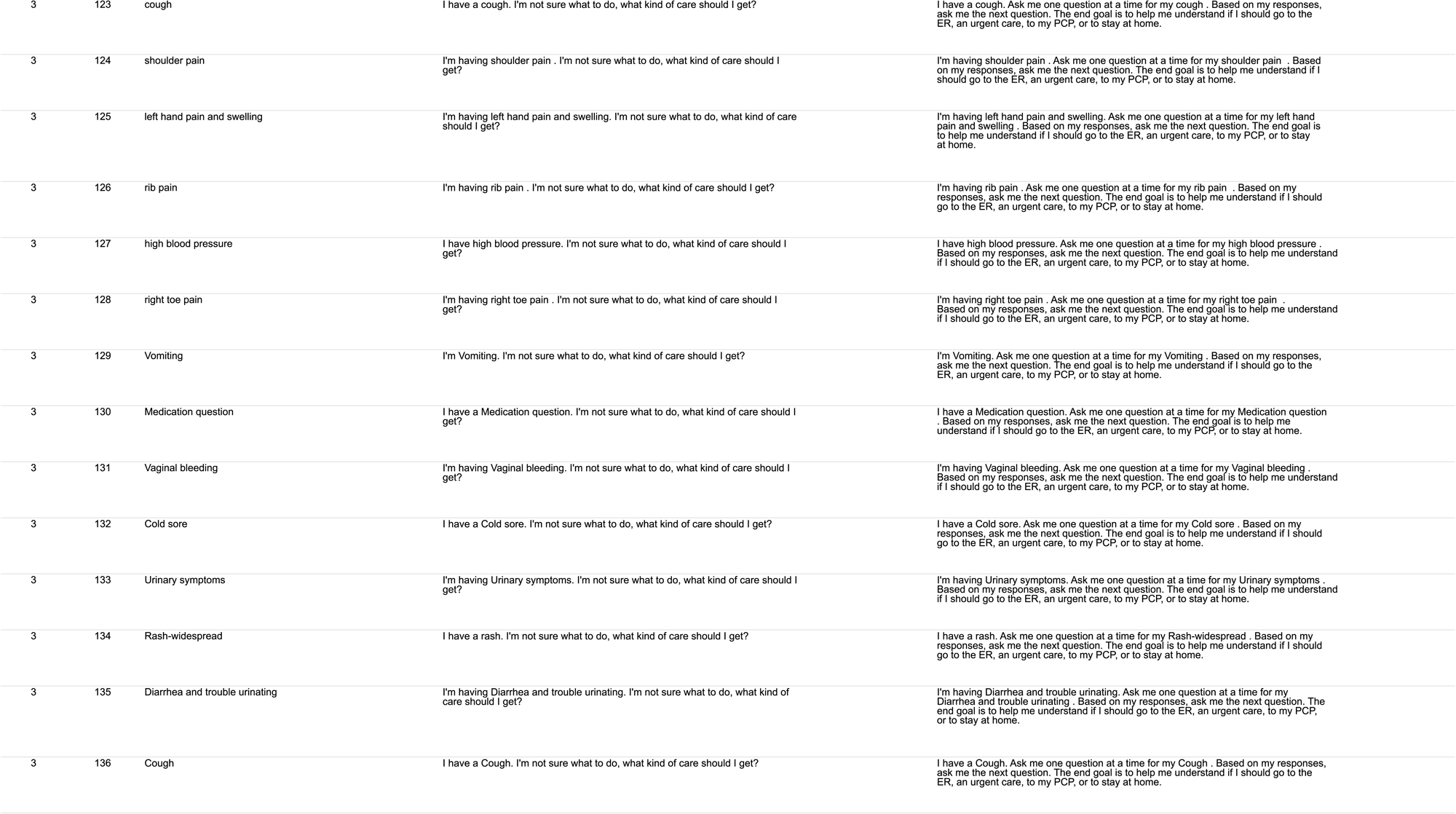
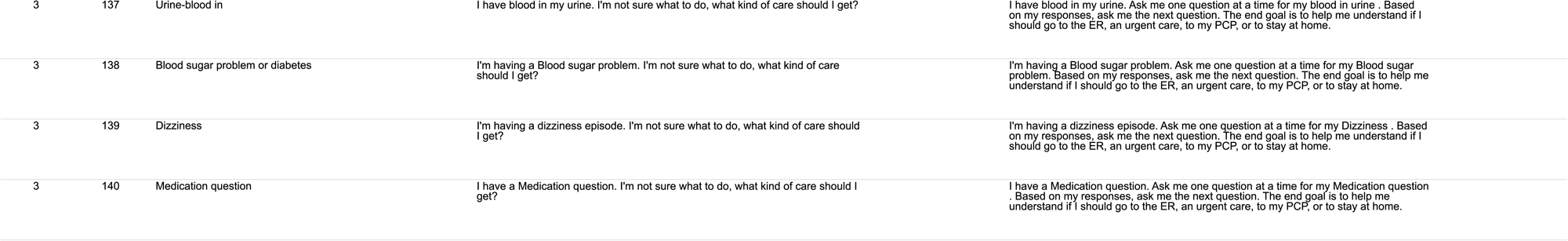
ChatGPT Health Prompts for dataset. All 255 cases are listed by dataset, prompt number, chief complaint and the natural and multi-turn prompt presented to ChatGPT Health. Dataset 1 utilized synthetic vignettes derived by clinicians. Dataset 2 and 3 prompts were derived from documented chief complaints and provided nurse-triage conversations.

